# A spatial single-cell transcriptomic atlas of metastatic breast cancer progression

**DOI:** 10.64898/2026.09.04.26361940

**Authors:** Zhanna Dauyey, Arno Velds, Roán Gobits, Iris de Rink, Karianne Schuurman, Merel Roest, Isabel Mayayo-Peralta, Suzanne Beerthuijzen, Marcos Da Silva Guimaraes, Sten Cornelissen, Joyce Sanders, Marja Nieuwland, Iris M. Seignette, Francesca Paolucci, J. Christoph Lampert, Carmen Jeronimo, Rui Henrique, Roderick Beijersbergen, Sebastian Gregoricchio, Wilbert Zwart

## Abstract

Metastatic breast cancer represents a major societal burden and is a main cause of cancer-related death in women, with limited rationale-based treatment options and often dismal outcomes. Intra-tumor heterogeneity, tumor-stromal interactions and clonal selection are considered to play a major role in metastasis formation and disease progression, but these processes remain incompletely understood.

Leveraging spatial transcriptomics as cost-effective method for generating transcriptomics data at single-cell resolution, we present a single-cell spatial transcriptomics atlas of 126 tumor samples from 44 metastatic breast cancer patients, encompassing 520,850 cells from primary and metastatic lesions, with 171,766 cancer cells and 349,084 cells belonging to the tumor microenvironment. For most patients, multiple time-points of sample collection throughout metastatic disease progression were analyzed, allowing for detection of treatment-induced transcriptional changes driving therapy resistance within the tumor cell compartment. Distinct immune cell subpopulations were enriched upon progression following specific therapeutics and were predictive for hormone receptor loss. By inferring DNA copy number alterations from the single-cell transcriptomics data, we tracked subclonal tumor cell selection upon progression and identified critical transcriptomic features of outcompeting clones with future metastatic potential.

Cumulatively, we present a unique single-cell spatial framework on metastatic breast cancer development and progression, in a highly complex systemic treatment landscape.

## Introduction

Breast cancer is the most-common malignancy in women, accounting for over 2.3 million women who are diagnosed with the disease worldwide, each year(1). Breast cancer is considered a heterogeneous disease, and primary tumors are classified over distinct molecular subtypes: luminal (A or B), HER2-amplified or basal-like/triple-negative (TNBC)(2). These breast cancer subtypes expose a clearly distinct clinical trajectory, with TNBC having most relapses and disease progression within the first 5 years after diagnosis (3). In contrast, luminal breast cancers are classically considered less aggressive, but can experience relapses for decades after initial diagnosis, with progression risk ultimately surpassing the other subtypes (4,5). Approximately 70% of breast cancer cases are of the luminal subtypes (A or B) and stain positive for Estrogen Receptor alpha (ERα) (6–9). For these tumors, ERα is considered a critical driver of growth and progression (10–12). Upon activation by its natural ligand estradiol, ERα serves as transcription factor and occupies the DNA to regulate responsive genes over large genomic distances, to drive genetic programs that sustain tumor cell proliferation and survival (13,14).

Overall, breast cancer is considered a well-treatable tumor type, with 5-year survival well over 95% after primary diagnosis (15). However, late relapses are often observed, especially for the luminal subtypes (16–18), and annually over 670,000 patients succumb to the consequences of the disease, rendering it the most-common cause of cancer-related death in women (1).

Over the past decades, large consortia-based efforts have genetically, epigenetically and transcriptionally profiled primary breast tumors, expanding our understanding of inter-tumor heterogeneity and deepening our knowledge on the genetic changes associated with the disease (19,20). Further highlighting the heterogeneous nature of the disease, prior single-cell studies in primary breast cancer have demonstrated distinct malignant states among tumor cells (21). Alongside diverse cancer subpopulations, stromal and immune cell types (such as cancer-associated fibroblasts (CAFs) and myeloid subtypes) have an impact on immune evasion and therapy response, and are associated with clinical outcome and endocrine therapy sensitivity (22). However, most prior single-cell studies (19–22) in breast cancer were focused on primary tumors, leaving progressive metastatic disease largely uncharacterized at the single-cell level.

More recently, extensive investments are being made in genomic profiling of advanced metastatic breast cancer (23,24). These and other studies identified distinct genetic alterations that drive endocrine-therapy resistance, including activating point mutations in the *ESR1* gene (encoding for ERα) (25), or a somatic loss of ERα expression (26,27); both of which selectively enriched after prolonged adjuvant endocrine treatment, but not to be attributed to the metastatic process itself. In comparing primary disease with metastatic lesions, but also between different metastatic lesions in different organs, other critical drivers such as HER2 status remained largely stable (28,29). In metastatic disease, transcriptomics analyses revealed a frequent switching to more aggressive subtypes, with luminal A converting in 50% of cases to the higher proliferating luminal B subtype, but transition to HER2 amplified or TNBC was also observed (23,30). Nevertheless, longitudinal analyses of metastatic breast cancer progression and sequential sampling of metastatic material, in relation to treatment and through the lens of intra-tumor heterogeneity and tumor microenvironment (TME) interplay, are currently lacking.

Despite the great majority of breast cancer deaths follow metastatic relapse under treatment (31), most of the current reference studies rely on primary tumors removed before therapy began. The effects on tumor cells and their microenvironment across successive lines of treatment has been inferred from multiple patients rather than observed within the same individual (32). In this study, we aimed to resolve how systemic therapy reshape metastatic breast cancer: how malignant cells rewire their transcriptional programs, which clones expand at metastatic sites, and how the immune and stromal compartments are remodeled in a treatment-specific manner. To address this question, we leveraged spatially resolved transcriptomics data as a cost-effective method to analyze 126 tumor samples at single cell resolution, from 44 breast cancer patients diagnosed with metastatic disease. Malignant pleural effusions, removed for palliative care and collected throughout disease progression, allowed for temporal analyses on tumor evolution, in response to treatment and in the context of a dynamic microenvironment. Our studies show treatment-induced reprogramming of the TME and tumor-intrinsic transcriptional reprogramming, in a highly treatment-specific fashion. Primary tumor TME composition had predictive value for loss of ERα expression and future metastatic potential, and copy-number variation (CNV) analyses revealed tumor-intrinsic features associated with metastatic outgrowth, treatment resistance, and the path of metastatic progression. These studies serve as framework and a rich resource for future translational studies on metastatic breast cancer evolution.

## Results

### Overview and single-cell atlas of metastatic breast cancer

To comprehensively study alterations in tumor cell composition and cellular signaling in metastatic breast cancer, we made use of the 10x Genomics Xenium spatial transcriptomics platform (**Fig. 1A**). Our study comprises a total of 126 tumor samples (20 primary tumors, 106 metastatic lesions) from 44 breast cancer patients (42 women, 2 men) with metastatic disease. Among these, 33 patients (75%) were diagnosed with ERα+ luminal breast cancer, 10 (22.7%) with triple-negative breast cancer (TNBC), and 1 (2.3%) with HER2+ disease (**Fig. 1B** and **Supplementary Table 1**). Metastatic material was collected and was suitable for spatial transcriptomics analysis for 42 of the 44 patients (**Fig. 1C** and **Supplementary Table 2**). A major strength of our study is that metastatic samples from multiple time points were available for 32 patients, with up to 10 samples (the primary lesion and 9 sequential malignant pleural effusion (MPE) samples) collected from patient M-29 (**Fig. 1C**). In addition to primary breast tumors (n=20), metastatic sites included skin (n=2), ovarian (n=4), recurrent breast tumors (n=4), breast angiosarcoma (n=1), lymph nodes (n=7) and pleural space metastases in form of MPE (n=84) (**Fig. 1C**). For one patient (M-36), samples were collected at autopsy, providing information from 7 distinct metastatic sites (**Fig. 1C**).

Due to the large number of paired metastatic lesions and detailed clinical data, our dataset represents a unique resource to study how disease progression and treatment in metastatic setting impacts metastatic cancer biology. For each patient, multiple tissue samples were included on a tissue microarray (TMA), with two 1 mm cores per sample resulting in 90 cores from 16 patients on average per slide. Each core contained 93–16,931 evaluable cells (**Supplementary Fig. 1A**) suitable for downstream analyses (see *Materials and Methods* section). Replicate core heterogeneity was estimated by performing Louvain clustering on the gene expression for a replicate set and averaging the lowest fraction of the normalized contribution across all clusters. Cores obtained from MPE samples show very little variation, derived from a relatively homogeneous suspension of the same cell pool (**Supplementary Fig. 1B**). In contrast, the solid tumors span a wider range of inter-core heterogeneity, likely attributed to intra-tumor heterogeneity.

A total of 520,850 individual cells were successfully detected using the Xenium platform (**Fig. 1D**). Most of the cells were identified in MPE samples (209,660: 40.3%), followed by primary breast cancer sites (125,815: 24.2%) and all the other metastatic sites (185,375: 35.5%) (**Supplementary Fig. 1D-E**). To better define the individual cell identities, we used the *spacexr* package (33) for cell type annotation, exploiting previously published single-cell RNA-seq data performed in human breast cancer samples (21) (**Fig. 1E** and **Supplementary Fig. 1E**). Across individual patients, the ratio of cancer to TME compartment, as well as the total number of cells analyzed, varied substantially (**Fig. 1F**). Overall, we found that 59.9% (n=312,139) of all cells belonged to the immune compartment, followed by cancer cells (33.0%, n=171,766) and non-immune stromal cells (7.1%, n=36,945) (**Supplementary Fig. 1E**).

On average, we were able to detect ∼221 genes per cell (median: 167) (**Supplementary Fig. 1F**), with a comparable number across tissue types (**Supplementary Fig. 1G**). Interestingly, the number of genes identified in cancer cells was slightly higher than in the other cell types (**Supplementary Fig. 1H-I**). Furthermore, variability in the number of genes detected was also observed across individual patients (**Supplementary Fig. 1J**).

As expected and consistent with previous reports (34–36), dimensional reduction analyses revealed overlap of distinct stromal cell populations across patients, whereas tumor cells segregated per patient (**Fig. 1D**). In line with these observations, uniform manifold approximation and projection (UMAP) of exclusively cancer cells (**Fig. 1G**, *left*) showed a segregation of these cells by patient (**Fig. 1G**, *middle*), yet remaining distinct between primary and metastatic sites (**Fig. 1G**, *right*). Altogether, these findings suggest that, while clear transcriptomic alterations occur between tumor cells at different stages, patient-specific features remain dominant.

Patients representing different breast cancer subtypes were included in the study. This was confirmed by single transcript levels for classical marker genes, including *ESR1*, *PGR*, *ERBB2*, as well as on pathway level (**Fig. 1H-I**). Expression patterns of *ESR1*, *PGR*, *ERBB2,* as identified by spatial single-cell transcriptomics analyses, were successfully confirmed by classic immunohistochemistry staining and pathological analyses (**Fig. 1J** and **Supplementary Table 3**). For all patients, relevant clinicopathological parameters, treatment history, and follow-up data were collected, providing a unique resource to study metastatic breast cancer progression and therapy resistance at single-cell spatial resolution (**Supplementary Table 1,4**).

**Figure 1.**
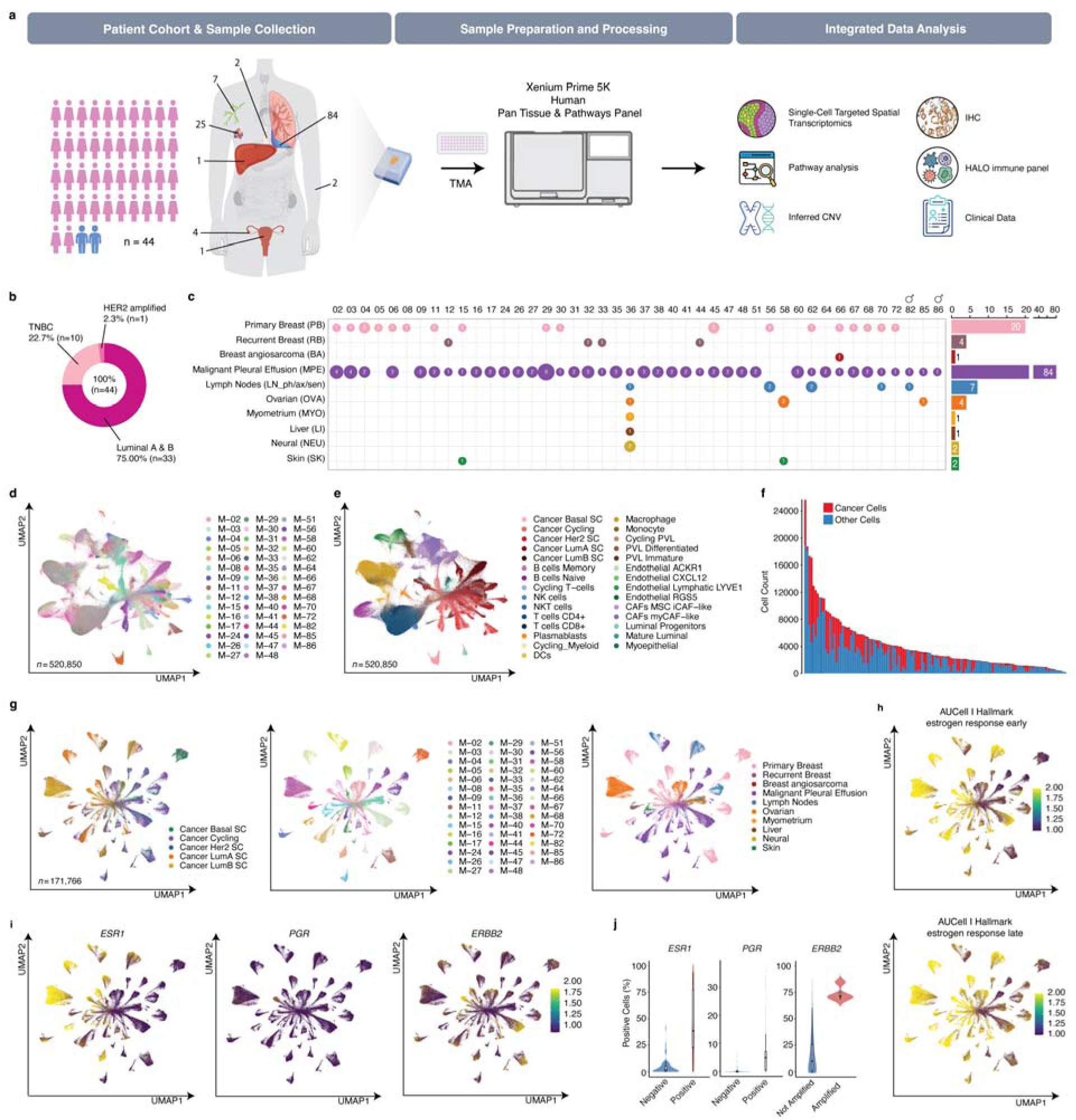
Overview and Single-Cell Atlas of Metastatic Breast Cancer. **a** Schematic workflow: study cohort, sample numbers from different metastatic sites, workflow and data types generated. In total 42 female and 2 male breast cancer patients were included. Numbers on the human figure indicate the number of samples from the corresponding metastatic site. Samples were multiplexed through tissue microarray (TMA)-based analyses, and data was analyzed on the 10x Genomics Xenium spatial transcriptomics platform. Data analysis types and validations are mentioned. **b** Donut chart depicting the distribution of breast cancer subtypes of primary lesion in the cohort. **c** Number of samples per tissue of origin per patient included for Xenium analysis. Bar plot (*right*) indicates total number of samples per tissue of origin. **d-e** UMAP projection of all cells (n = 520,850), colored by patient identifier (d) or by cell identity (e). **f** Total number of cells identified per patient sample, showing cancer (red) and non-cancerous (blue) cells. **g** UMAP projection of all cancer cells (n = 171,766) colored by breast cancer subtype (*left*), patient identifier (*middle*), and tissue of origin (*right*). **h** UMAP projection of all cancer cells. Color intensity depicts the expression signature scores for the Hallmark Estrogen response gene sets (early on *top*, late on the *bottom*). **i** UMAP projection of all cancer cells. Color intensity depicts the scaled level of expression of *ESR1* (*left*), *PGR* (*middle*) and *ERBB2* (*right*). **j** Violin and box plots showing the fraction of cells expressing *ESR1*, *PGR*, and *ERBB2*, classified by positive/negative (ER_α_, PR) or amplified/not amplified (HER2) according to immunohistochemistry status of the same samples (TMA slides). Boxplots indicate quartiles.

### Therapy-induced transcriptional reprogramming rewires metastatic landscape

In the treatment of metastatic breast cancer, therapy switching in response to disease progression is common practice, yet the rationale for selecting the most effective drug remains unclear. To identify commonalities and recurrent patterns across patients and different therapeutic regimens, we integrated our single-cell dataset with treatment history (**Fig. 2A**). In our cohort, patients received an average of six distinct therapies in the metastatic setting (**Fig. 2B**), with the total number of therapies ranging from 1 to 11 per patient (**Fig. 2B**). Patients in our cohort received multiple types of endocrine therapy, with aromatase inhibitors administered to 61.4% of patients (n=27), followed by tamoxifen (56.8%, n=25) and ERα degrader fulvestrant (29.5%, n=13). Targeted therapies encompass mTOR inhibitors (27.3%, n=12), CDK4/6 inhibitors (13.6%, n=6), and HER2 targeting drugs alone (13.6%, n=6) or as emtansine drug conjugate (4.5%, n=2). Cytotoxic chemotherapy compounds were widely prescribed, including antimetabolites (81.8%, n=36) interfering with nucleotide synthesis, taxanes (68.2%, n=30) disrupting microtubule dynamics, anthracyclines (65.9%, n=29), alkylating agents (54.5%, n=24) and platinum-based compounds (31.8%, n=14) that induce double-stand DNA breaks (**Supplementary Fig. 2A-B**). For a complete list of all prescribed therapeutics, see **Supplementary Table 4**.

Therapy switching typically occurs in response to clinical progression. At the metastatic stage of the disease, repeat sampling of the solid lesions is uncommon as this would give substantial discomfort without clear patient benefit, whereas sequential pleural drainage is often performed for palliative care. This renders MPEs a predominant source of metastatic cancer material in our study. Together with metastatic solid lesion samples, the sequentially collected MPE samples with variable intervals – ranging from a few days to several years – provide therefore a unique opportunity to explore mechanisms of therapy resistance by performing differential gene expression analyses between samples collected before and after a given treatment (**Fig. 2C, Supplementary Table 5**), allowed us to identify resistance mechanisms across a wide range of therapies (**Supplementary Table 6**). Focusing exclusively on the tumor cell population within our dataset, we performed over-representation analysis (ORA) on the differentially expressed genes between samples collected before and after progression on aromatase inhibitors (18 samples from 9 patients), tamoxifen (12 samples from 6 patients),

HER2 inhibitors (4 samples from 2 patients), DNA-damaging agents (16 samples from 8 patients) or antimetabolites (14 samples from 7 patients) (**Fig. 2C**). In patients progressing after aromatase inhibitors, hallmarks for ‘E2 response’, and ‘Reactive Oxygen Species’ were enriched, in line with previous studies (37–39) (**Fig. 2D**), while on a gene level, this was reflected by upregulation of *HSD17B1* (**Supplementary Fig. 2C**) (restoring local estrogen synthesis) (40), *NDRG1* and *GHITM* (associated with resistance to estrogen deprivation and stress response) (41–44), along with genes involved in apoptotic regulation such as *GSR* and *NQO1* (45,46) (**Fig. 2E**). These findings suggest that suppression of estrogen production exerts selective pressure under which tumor cells may adaptively rewire hormone signaling and survival programs or favor the proliferation of resistant sub-clones. Progression on antimetabolites was associated with enrichment of hypoxia and glycolysis pathways, indicating metabolic reprogramming as a potential resistance mechanism (**Fig. 2D**). For tamoxifen treatment, enrichment of the mTOR pathway after progression pointed to activation of this pro-survival and growth-promoting signaling cascade in tumor cells with an acquired resistance to the therapy (**Fig. 2D**). These transcriptomic observations were subsequently confirmed at the protein level using immunohistochemistry (**Fig. 2F**, *left*). Samples collected after progression on tamoxifen showed markedly elevated phosphorylated S6 (p-S6) ribosomal protein signal compared with pre-treatment samples (**Fig. 2F**, *right*), confirming mTOR pathway activation, in agreement with prior reports (47–49). Collectively, these findings demonstrate that metastatic breast cancer cells are subjected to therapy-associated evolutionary pressure while retaining patient-specific features, emphasizing both the complexity and the dynamic nature of metastatic progression.

**Figure 2.**
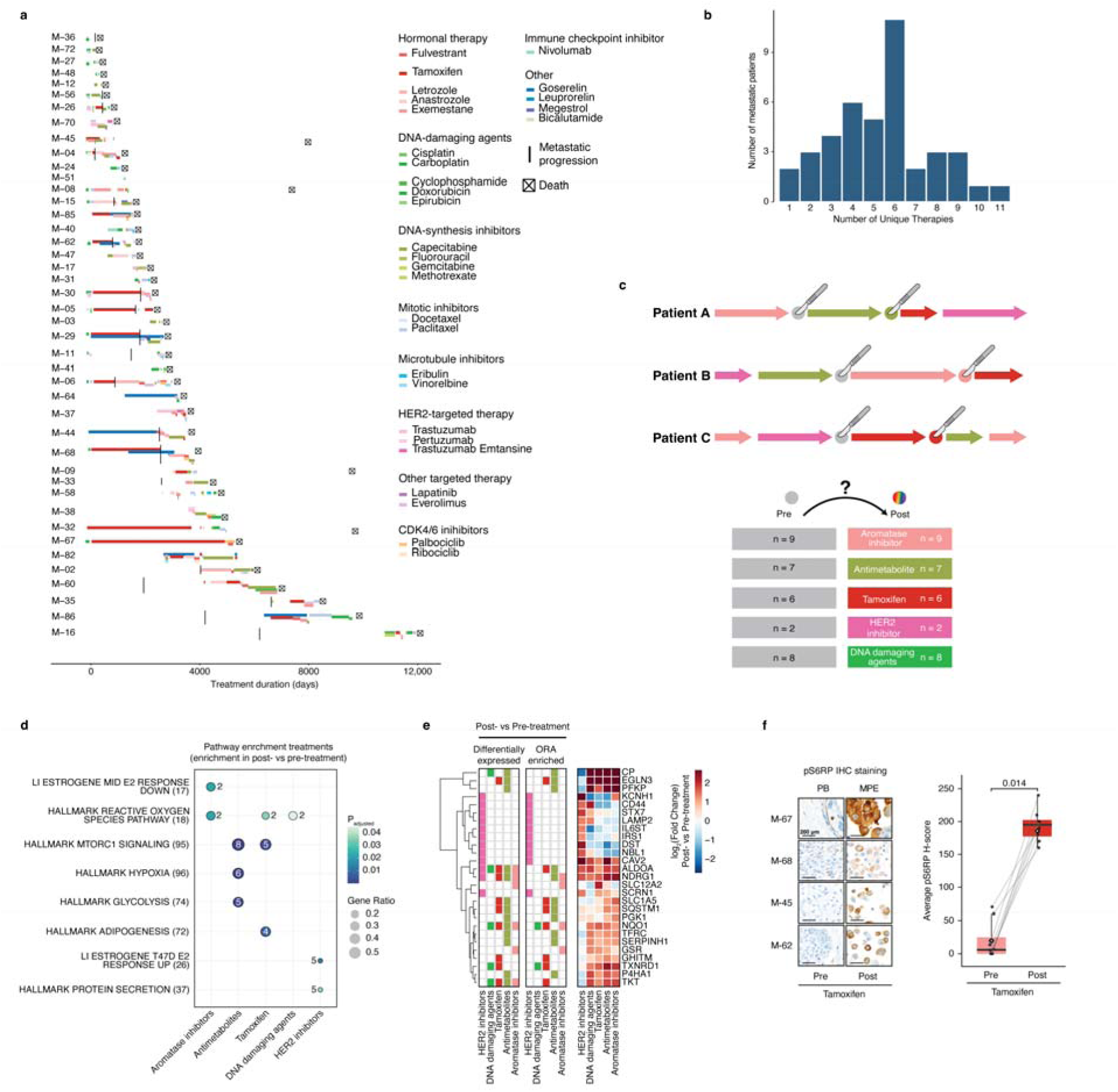
Treatment history and therapy-driven transcriptional reprogramming in metastatic lesions. **a** Comprehensive timeline, depicting the treatment history of the patient population, highlighting the administration of specific medications over time. Each horizontal track represents an individual patient (labeled M-## on the y-axis) over their total clinical course, with colored segments denoting the duration and sequencing of specific therapeutic interventions. Treatments are color-coded by pharmacological class. The x-axis indicates the treatment duration in days from the day of diagnosis. **b** Distribution of the number of unique treatments prescribed in the metastatic setting, for the patients in our cohort. **c** Schematic of pairwise comparison between tumor cells collected before and after specific treatments. By comparing samples collected before a treatment with samples after progression, transcriptomic alterations in response to therapy resistance are identified. **d** Combined graph of ORA on paired samples for tumor cells before and after progression on different therapies (aromatase inhibitors, antimetabolites, tamoxifen, DNA-damaging agents or HER2 inhibitors); genes upregulated in the post-treatment condition are analyzed. Pathway genesets are labeled by their nomenclature on the y-axis with the total number of genes in brackets. Dot size represents the gene ratio, the inner number corresponds to the number of the detected genes contributing to each specific pathway, while color intensity indicates significance (adjusted p-value, BH multiple testing correction). **e** Heatmap of log_2_(fold change) gene expression values for post-treatment comparisons across five therapeutic classes. Side-bar annotation (*left*) indicates genes reaching statistical significance for differential expression and ORA within each specific modality (colored by treatment group). Hierarchically clustering was performed using complete linkage based on euclidean distance. **f** Representative immunohistochemistry images (*right*) showing increased phospho-S6 ribosomal protein (pS6RP) signal in samples collected after progression on Tamoxifen compared with pre-treatment samples, with accompanying box plots of pS6RP H-scores (*left*) quantifying this increase. To determine statistical significance Wilcoxon signed-rank test (P = 0.014) was applied. Lines connect paired samples.

### Distinct therapies drive specific remodeling of the tumor microenvironment during metastatic progression

Tumor progression and therapy response are not solely determined by cancer cell-intrinsic features but are also critically affected by interactions with the surrounding stroma and immune cells (50). Therefore, we next studied the associations of treatment resistance, particularly in the metastatic setting, with the TME composition (**Fig. 3A**). In total, we identified 312,139 TME cells including B and T-cells, natural killer (NK) and NKT cells, myeloid cells, cancer-associated fibroblasts (CAFs) subsets such as myCAF-like cells (myofibroblastic CAFs, localized to invasive tumor regions (51), and MSC/iCAF-like cells (inflammatory CAFs, with cytokine-secreting and mesenchymal stem cell-like features (52)) and, endothelial and perivascular-like (PVL) cells (**Fig. 3B**).

In agreement with gene list-based cell type annotations, single-gene marker expression confirmed the identity of each stromal subset (**Fig. 3C**). Furthermore, spatial transcriptomics-based cell type identification was validated using multiplex-immunofluorescence HALO (Indica labs) imaging across selected samples for B cells, CD4⁺ T cells, Macrophages, employing well-established membrane markers specific to each stromal population (**Supplementary Fig. 3** and **Supplementary Table 3**). Finally, spectral flow cytometry (SFC) using a panel of 11 surface markers served as an orthogonal readout of sample composition (**Supplementary Fig. 4A,D**). The relative abundance of major cell populations was concordant with spatial transcriptomics, with modest shifts toward lower myeloid and higher T-cell proportions reported by SFC, potentially reflecting cryopreservation-induced biases (**Supplementary Fig. 4A**).

Having established the cellular identities and diversity of the stromal compartment, we next investigated how the relative abundance of these populations varied across patients, and in relation to the proportion of tumor cells within each sample. Ranking of the samples according to tumor cell percentage (TCP) over the entire cohort, revealed marked associations between tumor cell abundance and the surrounding stromal composition (**Fig. 3D-E** and **Supplementary Fig. 5**): macrophages (P = 9×10⁻, Mann-Whitney) and myeloid cells (P = 4.3×10^-7^, Mann-Whitney) positively correlated with high tumor cell content, suggesting a tumor-promoting, microenvironment, whereas CD4⁺ T cells displayed an inverse relationship (P = 6.7×10^-9^, Mann-Whitney). These findings suggest that higher tumor cell content is associated with reduced infiltration of helper T cells, consistent with previous reports of immune surveillance loss in more advanced lesions with dense tumor growth (53).

We then extended our stratification not only by tumor cell proportion (high versus low) but also across different tissue types, ERα levels and metastatic status (**Fig. 3F** and **Supplementary Table 7**). In metastatic breast cancer progression following adjuvant endocrine treatment, expression of *ESR1* is lost in approximately 22.5% of cases, a phenomenon known as “receptor conversion” (26). However, associations between receptor conversion and alterations in the TME remain largely unexplored. Across tissue types, we did not observe major differences in overall stromal composition (**Fig. 3F**). Importantly, within ERα-positive and ERα-converted MPE, all cancer cell subpopulations were more abundant in high-TCP samples compared with low-TCP samples, indicating that no single cancerous lineage drives this effect. In ERα-converted MPE, high-TCP samples showed strongest enrichment in highly proliferative (Cancer Cycling) and HER2-positive (Cancer HER2 SC) subpopulations. The overall immune and stromal composition of ERα-converted MPE remained largely similar to ERα-positive MPEs, indicating that TME retains luminal-like features despite ERα conversion rather than shifting toward a TNBC-like state (**Fig. 3F**). The only notable difference was the increase in CD4⁺ T cells in samples with low tumor cell content (|log_2_(fold change)| = 0.90, adjusted P = 0.04), indicating that helper T cells remain present in ERα-converted metastatic effusions, particularly in samples with lower tumor cell percentage (**Fig. 3F**).

To further explore the dynamic nature of the TME, we analyzed sequential samples from the same patients over time, alongside changes in their treatment regimens. This longitudinal analysis revealed substantial variability in TME composition over time (**Fig. 3G** and **Supplementary Fig. 6**), suggesting that systemic therapy and the development of treatment resistance may be linked to remodeling of the stromal and immune compartments. Consistent with this hypothesis, we observed distinct and reproducible alterations in specific stromal populations depending on the type of therapy administered: all subsets of cancer-associated fibroblasts (CAFs) and plasmablasts were consistently depleted following endocrine therapy, including both tamoxifen and aromatase inhibitors (MSC-like CAFs: AIs P = 0.002, tamoxifen P = 0.01; myCAF-like CAFs: AIs P = 6.10×10⁻, tamoxifen P = 0.002; plasmablasts: AIs P = 0.013, tamoxifen P = 0.002; paired Wilcoxon signed-rank test) (**Fig. 3H** and **Supplementary Fig. 7**). In contrast, cycling myeloid cells were enriched (P = 0.026, Mann-Whitney), and natural killer (NK) cells were decreased (P = 0.03, Mann-Whitney) specifically after aromatase inhibitor treatment (**Fig. 3H**). Standard DNA-damaging chemotherapy (including doxorubicin, cyclophosphamide, platinum-based agents; P = 0.016, Mann-Whitney) and tamoxifen (P = 0.01, Mann-Whitney) were associated with depletion of CD8⁺ T cells (**Fig. 3H**).

To assess whether the therapy-associated TME changes could be recapitulated *ex viv*o, we generated MicroOrganoSpheres (MOS) (54,55) from viably cryopreserved tumor samples. SFC confirmed that cell-type proportions of the MOS were comparable to the source tissues (**Supplementary Fig. 4B, 4D**). After 4-day treatment with carboplatin or tamoxifen, TME composition remained largely similar across conditions within each patient-derived model (**Supplementary Fig. 4C**). Together, these findings highlight therapy-specific modulation of TME composition that appears to be driven by context-dependent *in vivo* mechanisms associated with acquired therapy resistance rather than short-term, direct therapeutic effects on immune and stromal populations.

Having established that systemic therapies induce distinct and reproducible changes in the tumor microenvironment, we next wondered whether intrinsic tumor features might also shape immune composition independently of treatment. Remarkably, in primary tumors that later gave rise to ERα-converted metastatic lesions, B cells were significantly lower, affecting both naïve and memory populations rather than a single subpopulation (**Fig. 3I**, *top*). In the corresponding ERα-converted metastatic lesions, NK and NKT cells were enriched, indicating that receptor conversion is associated with remodeling of the innate immune compartment upon metastatic progression (**Fig. 3I**, *bottom*).

**Figure 3.**
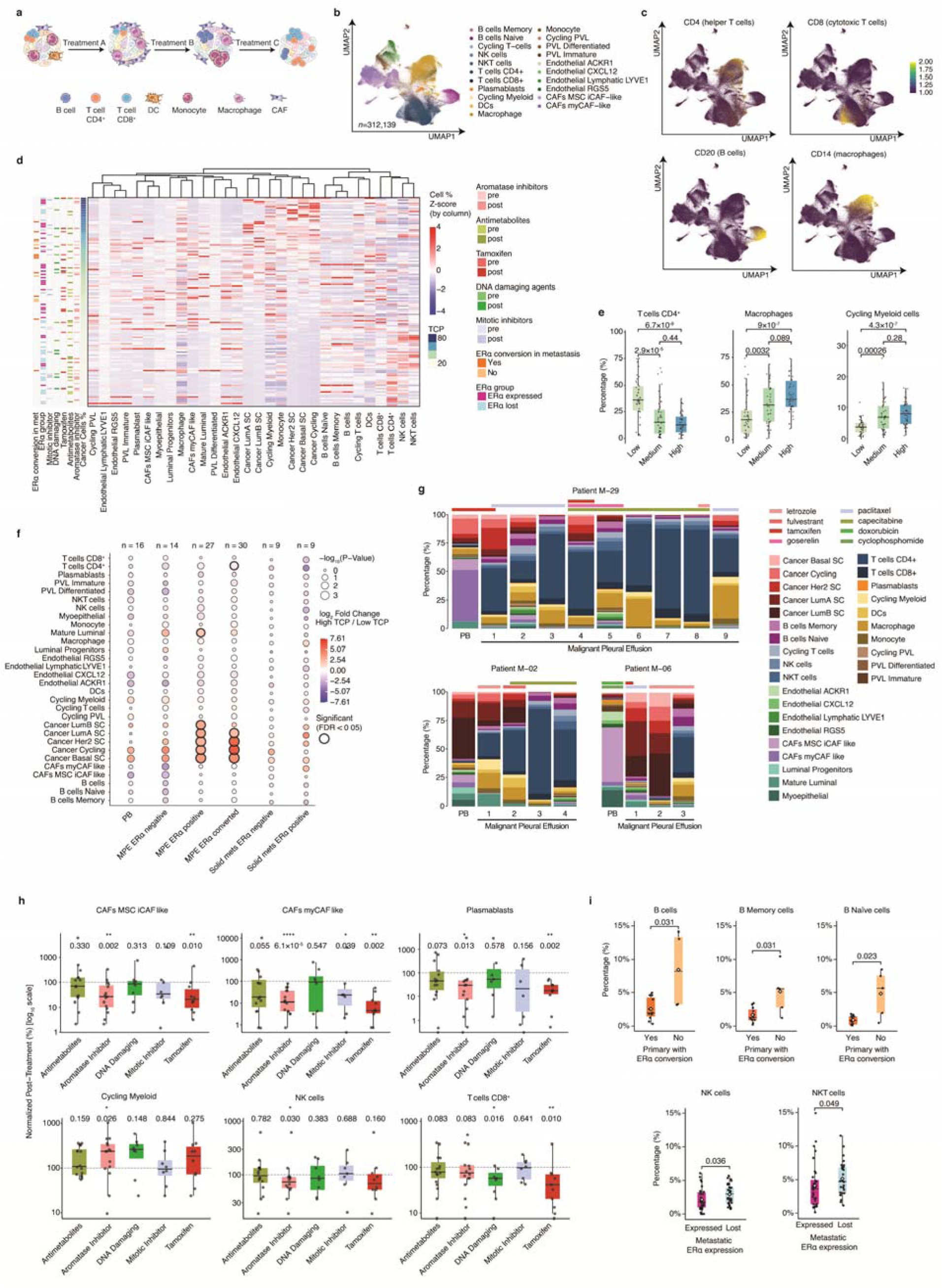
Remodeling of the tumor microenvironment across therapies and metastatic progression. **a** Schematic illustrating the relevance of the tumor microenvironment (TME) in metastatic disease and its modulation by different treatments. **b** UMAP projection of 312,139 TME cells classified using curated transcriptomic signatures, showing B cells, CD4⁺ and CD8⁺ T cells, NK and NKT cells, dendritic cells, monocytes, macrophages, multiple CAF subsets (myCAF-like, iCAF-like, MSC-like), endothelial subtypes (RGS5⁺, LYVE1⁺, CXCL12⁺, ACKR1⁺), and perivascular-like cells (immature, differentiated, cycling). **c** UMAP projection of all TME cells (n=312,139). Color intensity depicts the scaled level of expression of *CD4*, *CD8 (CD8A)*, *CD20 (MS4A1)*, and *CD14*. **d** Heatmap derived from the relative proportions of tumor, immune and stromal cell populations across all samples. Cell percentages (expressed as a percentage of total cells for cancer and total immune cells, and as a percentage of total immune cells for individual immune subpopulations) were scaled by column (z-score). Rows represent individual samples, ranked by tumor cell percentage (TCP), and columns are the annotated cell types that were hierarchically clustered using complete linkage based on euclidean distance. Row annotations indicate cancer cell content and clinical features with the following color encodings: by treatment, with lighter shades indicating pre-treatment status and darker shades, representing post-treatment status across five drug classes: Aromatase inhibitors (pink), Antimetabolites (olive), Tamoxifen (red), DNA damaging agents (green), and Mitotic inhibitors (blue); Primary tumors with ER conversion during metastasis were highlighted in orange, while stable ER status was shown in peach, and ER status of the metastatic sample indicated in magenta (ER expressed) and light blue (ER lost). **e** Boxplots showing percentages of CD4⁺ T cells (P = 6.7 × 10⁻^9^), macrophages (P = 9 × 10⁻^7^), and cycling myeloid cells (P = 4.3 × 10⁻^7^) across low, medium, and high tumor cell percentage (TCP) groups. Cancer cells and total immune cells are expressed as a percentage of total cells, whereas individual immune subpopulations are expressed as a percentage of total immune cells. Stratification of the samples was performed computing tertiles of the TCP distribution. Mann-Whitney U-test was applied. **f** The dot plot illustrates the differential abundance of 30 cell types across six clinical strata (Primary (PB), MPE ER negative/positive/converted, and Solid metastases ER negative/positive), with sample sizes (n) indicated for each group. The Y-axis shows specific immune, stromal, and cancer cell populations, while the X-axis categorizes these by tissue origin and ERα status. Statistical significance is represented by dot size, corresponding to the −log_10_ (Mann-Whitney U-test P-value). The color gradient denotes the log_2_(fold change) (High TCP / Low TCP). Points reaching statistical significance after Benjamini-Hochberg correction (FDR < 0.05) are highlighted with a thickened black border stroke. **g** Stackplots showing the longitudinal proportional changes of tumor and stromal populations in primary breast tumors and over sequential MPE samples and associated treatments. Bars are annotated with the specific therapeutic agents administered at each time point. **h** Boxplots of matched pre/post-treatment samples showing therapy-induced TME changes after treatments (antimetabolites, aromatase inhibitors, DNA-damaging agents, mitotic inhibitors, and tamoxifen). Cell type percentages (expressed as a percentage of total cells for cancer and total immune cells, and as a percentage of total immune cells for individual immune subpopulations) were normalized to the pre-treatment baseline ((post / pre) × 100; dashed line at 100%) and plotted on a log_10_ scale. Values above or below 100% show increased or decreased relative abundance for a cell type, respectively. Boxes show the median and interquartile range with 1.5×IQR whiskers; gray points represent individual samples. To determine statistical significance between paired pre-and post-treatment samples, Wilcoxon signed-rank test was applied. P-values are reported above each treatment group, with stars denoting levels of significance (^∗^p<0.05, ^∗∗^p<0.01, ^∗∗∗∗^p<0.0001). **i** Boxplots showing percentages of B cells (P=0.031), B Memory cells (P=0.031), and B Naïve cells (P=0.023) in primary tumors that gave rise to ERα-converted metastases (Yes) compared to those that did not (No) (*top*). Cancer cells and total immune cells are expressed as a percentage of total cells, whereas individual immune subpopulations are expressed as a percentage of total immune cells. On the *bottom*, NK cells and NKT cells in metastatic samples grouped by ERα expression (expressed vs. lost). Dots represent individual patient samples, while diamonds indicate group means. Statistical significance was determined using Mann-Whitney U-test.

### Defining clonal architecture and molecular determinants of metastatic potential

Following characterization of the cellular and immune landscape across primary and metastatic lesions, we next turned to the genetic basis of metastatic progression. Clonal selection is a well-established driver of resistance to systemic therapy (**Fig. 4A**) (56). To investigate the role of clonal selection in metastasis formation and therapy resistance, we inferred single-cell copy-number variation (CNV) profiles from our spatial transcriptomics data using *inferCNV* (57) and applied Louvain clustering to the resulting gene/cell copy number matrix to identify clonal groups (see *Materials and Methods* section). The accuracy of our CNV inference was validated in a subset of samples for which we previously generated Hi-C data (58) (**Supplementary Fig. 8**). As Hi-C is a bulk 3D conformation capture technique, we used these data to validate CNVs identified across all clusters within each corresponding sample (**Supplementary Fig. 8**).

In patient M-02, CNV clustering analysis revealed two genetically distinct putative subclones within the primary tumor (**Fig. 4B**). While Clone #1 (CNV clusters 0+1+5) was present in the metastatic lesions only at low frequency and progressively lost over time, Clone #2 (CNV clusters 2+3+6) metastasized and expanded to a distant site – at the expense of Clone #1 – being the predominant clone detected in MPE samples collected 11 and 14 years after primary diagnosis. ORA on differentially expressed genes (**Supplementary Table 6**) between the two clones in the primary tumor revealed distinct pathway activity profiles: the metastasizing Clone #2 exhibited downregulation of ERα response signatures and upregulation of mTOR and MYC hallmark pathways (**Fig. 4C**). Consistent with these findings, *ESR1* was readily expressed in the non-metastasizing primary Clone #1 as shown by spatial transcriptomics (**Fig. 4D**) and confirmed by immunohistochemistry (**Fig. 4E**), very low levels of *ESR1* were detected for the metastasizing primary Clone #2 and MPE samples (**Fig. 4D-E**). In patient M-68, we identified a metastasizing clone corresponding to inferred CNV cluster 2, which was not confined to a specific core within the primary tumor, but widespread across multiple regions of tumor tissue (**Fig. 4F**). ORA, comparing each cluster against all the other clusters (**Supplementary Table 6**), revealed all estrogen response signatures, including Hallmark for early and late estrogen response pathways (59) as well as Estrogen response signatures defined by Li *et al.* (60) to be upregulated in Cluster 2 (**Fig. 4G**). Visualization of spatial expression at both the pathway level (**Fig. 4H**) and the single-gene level (**Fig. 4I**, *left*) confirmed sustained ERα signaling at the transcript level specifically in the area corresponding to metastasizing clone, which was again validated by immunohistochemistry in both primary tumor cores and MPE samples (**Fig. 4I**, *right*).

In both cases, clonal expansion occurred following endocrine therapy, suggesting that while therapy may select for ERα-independent subclones from heterogeneous primary lesions, ERα-positive clones can also persist and expand despite treatment.

**Figure 4.**
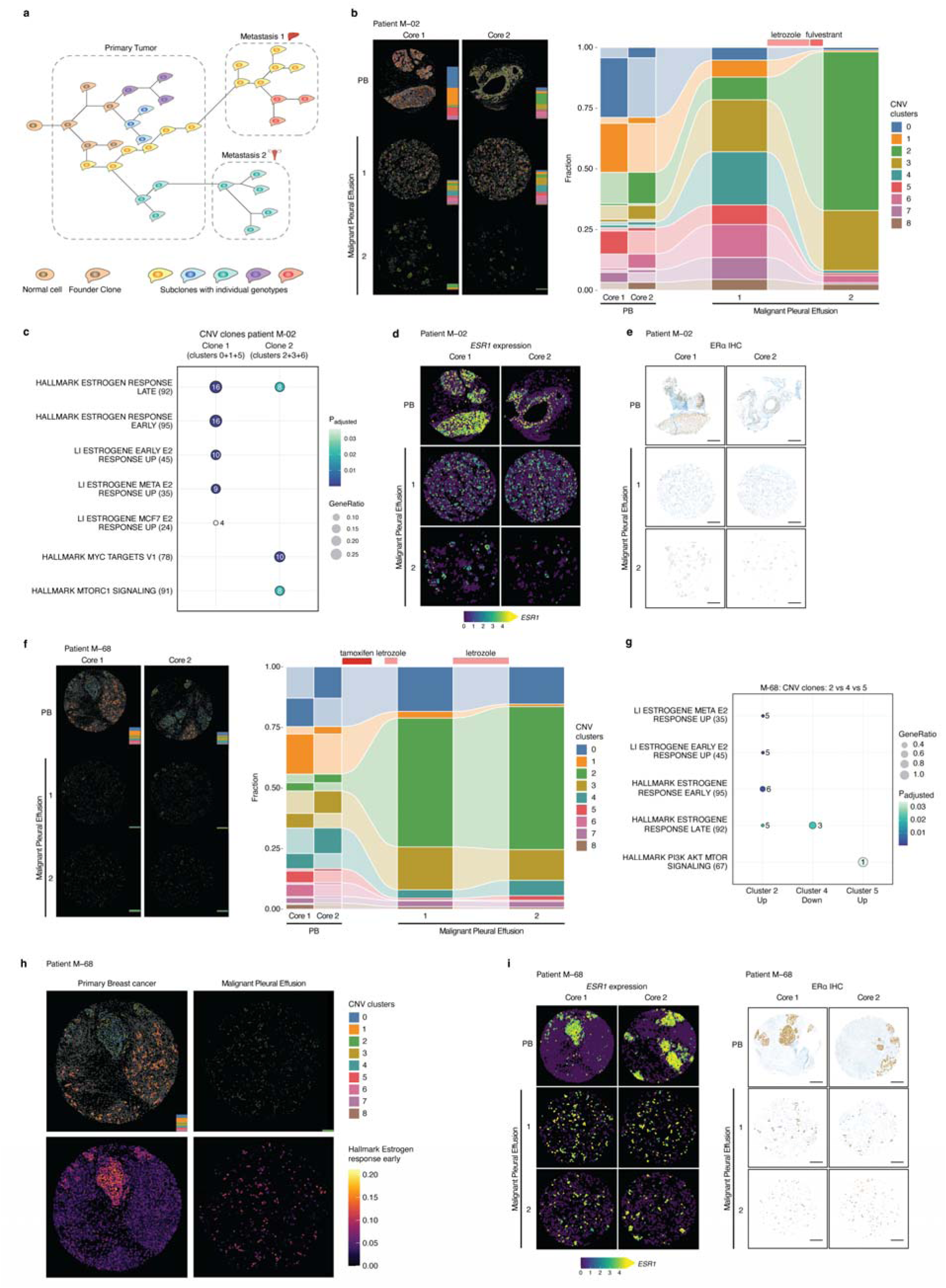
Clonal architecture and molecular features of metastasis-capable tumor populations. **a** Schematic cartoon illustrating the concept of clonal diversification and expansion during tumor progression. **b** Spatial visualization of Copy Number Variation (CNV) clusters 0-8 across the primary breast (PB) tumor and Malignant Pleural Effusion (MPE) sites 1 and 2 in patient M-02 (*left*). Alluvial plot quantifying the longitudinal change in the fractional abundance of CNV clusters from the PB through sequential MPE samples after endocrine therapy (*right*). **c** ORA of differential expressed genes between CNV clones 0, 1, and 5 against clones 2, 3, and 6 of patient M-02. X-axis categorizes up- or down-regulated pathways. Pathways are labeled by their nomenclature on the y-axis with the number of genes in brackets. Dot size represents GeneRatio and the number corresponds to the number of the detected genes contributing to each specific pathway. Color scale indicates P-adjusted values (BH multiple testing correction). **d** Spatial transcriptomic maps showing the expression levels of *ESR1* in the PB and sequential MPE samples of patient M-02. **e** Histological sections stained for ERα protein via immunohistochemistry (IHC) in the PB and sequential MPE samples, corresponding to the spatial transcriptomic regions in panel (d). Magnification scale is indicated by a black bar corresponding to 200μM. **f** Spatial mapping of CNV clusters in PB and MPE samples in patient M-68 (*left*). Alluvial plot showing the fractional distribution of CNV clusters 0-8 for the PB and post endocrine therapy for MPE samples 1 and 2 (*right*). **g** ORA of differential expressed genes between CNV clusters 2, 4, and 5 in patient M-68. Pathways are labeled by their nomenclature on the y-axis with the total number of genes in brackets. Dot size represents GeneRatio and the number indicates the number of the detected genes contributing to each specific pathway. Color scale indicates P-adjusted significance (BH multiple testing correction). Note that only differential expression list showing at least one enriched dataset are shown in the plot. **h** High-magnification spatial maps for patient M-68. *Left*: CNV cluster distribution in a PB core. *Middle*: Corresponding “Hallmark Estrogen Response Early” pathway scores in the same PB core. *Right*: “Hallmark Estrogen Response Early” pathway scores in MPE 1. **i** Spatial transcriptomic visualization of *ESR1* expression levels (*left*). Corresponding IHC staining for ERα protein across the PB and MPE samples 1 and 2 (*right*). Magnification scale is indicated by a black bar corresponding to 200μM.

### Reconstruction of metastatic trajectories in individual patients

In our cohort, there is one particular case for which we have secured samples from multiple metastatic sites (patient M-36; **Fig. 1C** and **Fig. 5A**), obtained in the context of a post-mortem resection. No primary tumor was available for this patient. Using the available metastatic samples, we explored whether our spatial transcriptomics dataset could be informative to extract the trajectory of metastasis formation within the patient. For this, we first attempted to use the inferred CNV signatures to identify any clonality expansion (**Supplementary Fig. 9A-B**). Unfortunately, this approach did not highlight any clear metastatic trajectory, as subclonal CNV variations between metastatic sites were modest. Moreover, analysis of cell type composition of the different metastatic sites was also inconclusive due to the limited variation on cell population ratios across samples (**Supplementary Fig. 9C**).

Therefore, we used *slingshot* (61) to infer pseudotime trajectories from gene expression patterns. We first benchmarked this approach in a sample (patient M-15) for which the temporal order of lesion occurrence is known (Primary tumor > Skin metastasis > MPE; **Supplementary Fig. 9D**). We were able to identify two distinct trajectories indicating that, consistent with clinical history, both metastases originated directly from the primary breast tumor (**Supplementary Fig. 9E**). We then employed *tradeSeq* (62) to identify the genes mostly contributing to the identity of each trajectory (**Supplementary Fig. 9F**). ORA of the top 50 genes showed that the transition from primary tumor to skin metastasis is characterized by an enrichment of mTOR signaling genes (**Supplementary Fig. 9G**).

We next applied the same approach to the patient M-36 (post-mortem resection case) and identified four trajectories with a common origin represented by the ovarian metastatic lesion (**Fig. 5A-B**). This observation suggests the occurrence of multiple cycles of dedifferentiation and phenotypic adaptation, rather than linear clonal succession. For each trajectory, we selected the top 50 driving genes (**Fig. 5C**) and performed ORA analyses (**Fig. 5D**). The latter highlighted that Trajectory 1 is enriched for genes regulated by MYC, while Trajectory 4 is characterized by E2F target genes (**Fig. 5D**).

Altogether, we showed that spatial transcriptomics data can provide insights into the temporal order of the cancer progression and highlights distinct parallel trajectories in metastatic spreading.

**Figure 5.**
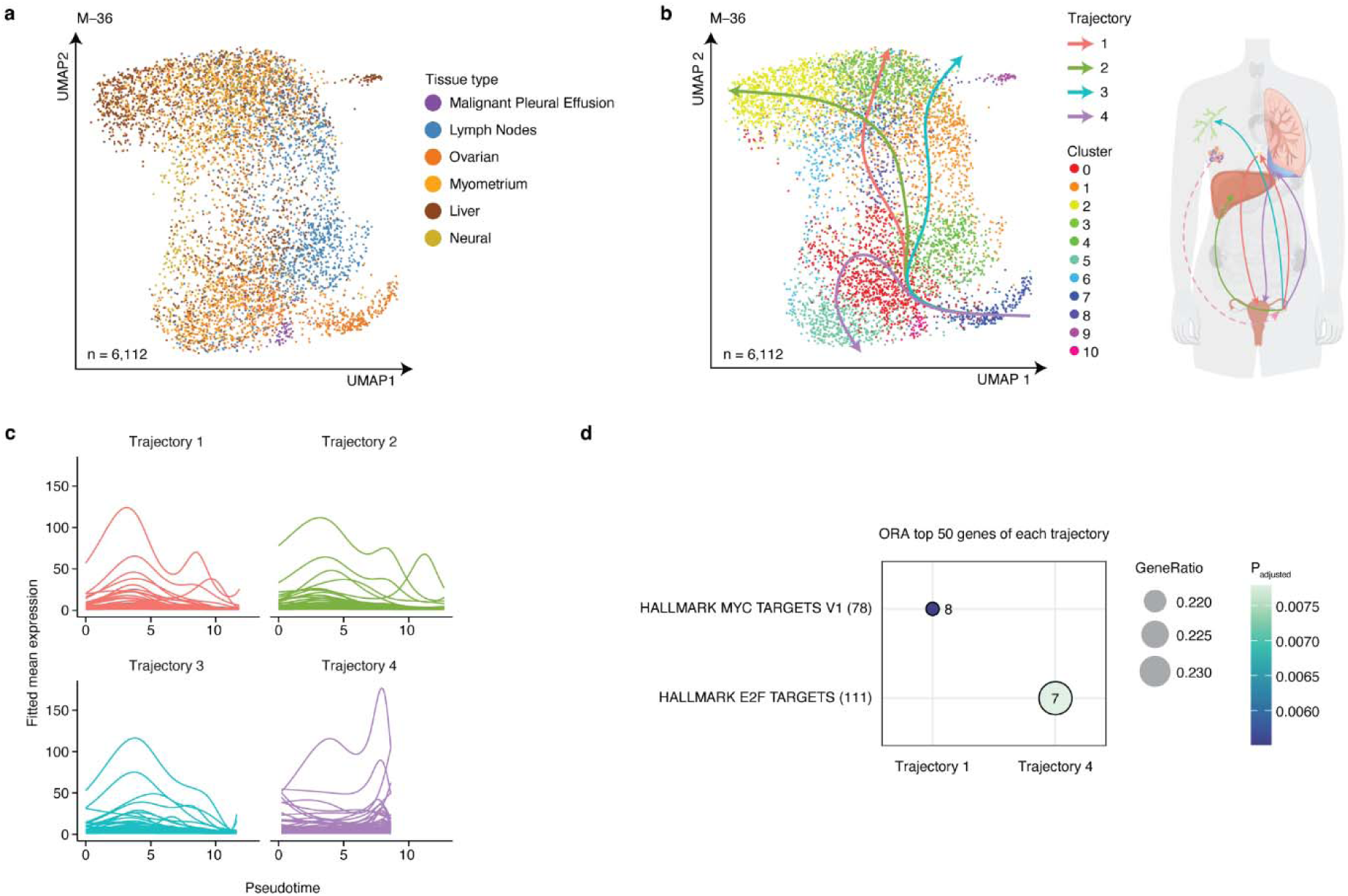
Reconstruction of metastatic events using pseudotime trajectories. **a** UMAP projection of all cancer cells belonging to patient M-36 (n = 6,112), colored by patient lesion location. **b** On the *left*, UMAP projection of all cells belonging to patient M-36 (n = 6,112), colored by *Seurat* defined expression clusters. Arrows indicated pseudotime inferred trajectories. The same trajectories are represented on the *right* with arrows connecting different organs, reflecting the inferred order of the metastatic lesion events. **c** Fitted expression pattern of the top 50 genes that most characterize each trajectory defined in (b). **d** ORA of the top 50 genes in (c). Pathways are labeled by their nomenclature on the y-axis with the total number of genes in brackets. Dot size represents GeneRatio and the number indicates the number of the detected genes contributing to each specific pathway. Color scale indicates P-adjusted significance (BH multiple testing correction). Note that only trajectories showing at least one enriched dataset are shown in the plot.

## Discussion

While most functional genomics studies in breast cancer have been focused on primary tumors, our understanding of metastatic disease progression and treatment-induced alterations in tumor biology is lagging behind. In this study, we provide a comprehensive single cell spatial analysis of metastatic breast cancer progression over time, using sequentially collected specimens from the same patient. A critical aspect in our study design is the collection and analysis of MPEs. Autopsy studies (63,64) demonstrate pleural involvement in 50-80% of patients with metastatic breast cancer, of which about 30% require drainage for symptom relief (63,65). These routine drainages provide sufficient metastatic material throughout disease progression as a part of palliative care, unlike metastatic biopsies, which are often low-yield, highly invasive with substantial burden on the patient, and rarely allow for longitudinal sampling. Such repeated sampling is essential to better understand the dynamic nature of cancer progression and tumor adaptation to drug-induced selection pressure remain poorly characterized largely due to limitations of metastatic biopsies. MPEs exhibit similar cancer subtypes (28,66), mutation profiles (67), gene expression profiles (68) to other metastatic sites, and biomarkers identified in MPEs are prognostic across multiple types of cancer (69,70), positioning their use as representative of progressive metastatic disease rather than a distinct entity.

While preclinical studies have established that prior treatments can reshape the tumor biology and reprogram cancer driver dependencies, clinically such observations have been mostly limited to mutational analyses in non-hematological malignancies (71–74). Our longitudinal analyses of breast cancer progression is innovative on a number of topics: transcriptional programs that emerge upon resistance to a defined therapy in metastatic disease, distinct alternations in stromal cell composition after disease progression, stromal features in primary lesions that can predict later conversion of estrogen receptor expression status upon metastatic dissemination and the selective metastatic potential of specific clones within primary breast tumors.

Neoadjuvant studies have demonstrated that therapies can induce distinct transcriptional programs, but these efforts have been limited in scope and restricted to only a few treatments (75–77). Here, we expand this understanding by systematically defining therapy-induced transcriptional alterations in metastatic lesions, separating out tumor-cell–intrinsic from tumor-cell–extrinsic features. This approach allows the identification of treatment-induced novel vulnerabilities, such as mTOR pathway activation emerging upon tamoxifen exposure, that may be considered of interest to pursue in future clinical studies.

Beyond tumor-intrinsic features, our analyses showed coordinated therapy-associated remodeling of the tumor microenvironment. These observations suggest that distinct treatment regimens can differentially reshape immune and stromal populations, potentially contributing to therapy resistance or altered anti-tumor immunity. Prior studies have reported elements of these responses in isolated contexts, such as CAF contributions to endocrine resistance (78–80) and chemotherapy-induced activation of cytotoxic T cells (81,82), but these observations have been fragmented, focused on single treatments, and rarely examined during metastatic progression. In contrast, our longitudinal profiling demonstrates how multiple therapies collectively reshape both tumor-intrinsic transcriptional states and tumor-extrinsic microenvironmental features over the course of disease progression. Interestingly, immune profiles of ERα-positive primary tumors appeared predictive of the ultimate development of receptor converted ERα-negative metastatic lesions, exposing decreased B-cell infiltration. In contrast, metastatic lesions show enrichment of innate immune populations, including NK and NKT cells, following loss of ERα expression. These observations highlight the complex interplay between tumors and surrounding immune cells, jointly imposing selective pressures to shape evolving tumor states.

For a subset of patients in our cohort, we observed genetically distinct clones within the primary lesion with selective capacity for metastatic dissemination. These observations highlight the necessity of extensive pathological interrogation of the primary lesion and raise caution when interpreting small sections of the primary tumor (e.g., through biopsy procedures), which may not capture the full clonal diversity. In some cases, adjuvant endocrine therapy coincided with the expansion of specific clones, yet the biological consequences of treatment selection highly diverged, selecting for ERα-low tumor clones or contrarily persisting ERα action. These observations highlight the complexity of therapy-driven selection and indicate that endocrine therapy imposes a common selective pressure, but that tumors achieve adaptation through diverse, patient-specific evolutionary routes.

Inferred CNV analysis of tumor cells and comparative visualization of TME cell-type composition of multiple metastatic sites from the same patient revealed limited clonal diversity and TME changes. These observations suggest that metastatic site-specific microenvironmental cues have limited impact on distinct (sub)clonal selection, and that treatment-induced alterations in tumor biology appear the most-dominant feature of genetic drift and clonal selection. Future studies should be designed to identify distinct (epi)genomic features that define the clone at-risk-of-metastasis, to enable tailoring of therapeutic strategies accordingly.

By transcriptionally profiling clinically well-annotated metastatic breast cancer throughout disease progression at single cell resolution, we provide an extensive resource that allows for in-depth investigation of therapy-induced tumor-intrinsic and -extrinsic features, clonal evolution, and metastatic trajectories. As a resource, our study supports the integration with multi-omic datasets, validation of predictive biomarkers, and development of models to anticipate treatment response and guide precision therapeutic strategies for metastatic breast cancer patients.

Even though spatial transcriptomics data was generated, the implementation of the spatial data itself in our analyses was modest, and mostly limited to clonality analyses (Figure 4). Despite extensive attempts, no clear patterns on cellular proximity, neighbor niches or cell-cell interactions were observed, in relation to treatment history, therapy response or subtype. Critically, this technology allowed us to analyze over half million cells from 126 tumor samples in a highly cost-effective manner, to reveal novel associations of microenvironmental factors with tumor hormone receptor conversion and treatment response, along with the possibility to track clonal evolution over time. A second limitation in our study, is the relative enrichment of luminal tumors relative to the other subtypes, in part due to a pleura-selective organotropism of this subtype (83). Future studies should be defined to allow for a better representation of less-prominent subtypes, including basal and HER+ positive tumors.

Cumulatively, our dataset represents a powerful resource for future studies on metastatic progression, specific treatment-induced changes on gene expression, cancer and microenvironment cell composition.

## Data availability

Raw and analyzed single-cell spatial transcriptomics and spectral flow cytometry data are available at the BioImage Archive under accession number S-BIAD2706. The data can also be explored at the Single Cell Portal under accession number [SCP3851](https://singlecell.broadinstitute.org/single_cell/reviewer_access/7ea63c58-e239-433c-9fa0-283848f9ff4f).

All the codes used for the analyses presented in this manuscript are available on GitHub at https://github.com/sebastian-gregoricchio/Dauyey_Velds__BRCa_single_cell_transcriptomic_atlas_CODES.

## Author contributions

Conceptualization: Z.D., A.V., S.G. and W.Z. Methodology: Z.D., A.V., M.N., S.C., I.M.S., R.B., F.P. and S.G. Software and Formal Analysis: A.V., S.G., Z.D., and I.d.R. Investigation: Z.D., A.V., M.N., S.C., I.M.S., R.G., and S.G. Pathological Assessment: M.D.S.G. and J.S. Resources: K.S., M.R., I.M.P., S.B., J.C.L., C.J., R.H., R.B., and W.Z. Data Curation: A.V., I.d.R., Z.D., and S.G. Supervision: S.G. and W.Z. Project Funding and Administration: W.Z. Z.D., A.V, S.G. and W.Z. wrote the manuscript, with input from all co-authors. All authors read and approved the final manuscript.

## Acknowledgements

We thank members of the Zwart and Bergman labs for valuable feedback, suggestions and input. We thank Elzo de Wit and Teun van den Brand for their support in the Hi-C data analyses. We would like to thank Nuno Padrão for his essential contribution in collecting malignant pleural fluid samples for this study. We would like to acknowledge the Research High Performance Computing (RHPC) facility of the Netherlands Cancer Institute (NKI) to have enabled us to perform all the computations required to analyze the data generated, the NKI Genomics Core Facility (GCF) for spatial transcriptomics data generation and bioinformatics support, the NKI-AVL Core Facility Molecular Pathology and Biobanking (CFMPB) for patient samples handling.

## Declarations

### Ethics approval and consent to participate

This study was approved by the local medical ethics committee of the Netherlands Cancer Institute and Portuguese Oncology Institute of Porto (Institutional Review Board (IRB) reference number: CFMPB411) and complies with the ethical principles of the Declaration of Helsinki. All patients provided informed consent for translational studies.

## Material and Methods

### Patient sample collection

Formalin-fixed paraffin embedded (FFPE) primary and metastatic breast cancer specimens were collected from the NKI Biobank. Malignant pleural effusion samples were collected between 2013 and 2020, at the Netherlands Cancer Institute and at the Portuguese Oncology Institute. Directly after drainage, pleural effusion fluid was centrifuged at 1600 g for 8 min, the resulting cell pellet was resuspended in 25 mL of erythrocyte lysis buffer (5mM KHCO_3_, 75mM NH_4_Cl, 500mM EDTA) for 10 min at room temperature (RT). Following lysis, the buffer was removed, and the cells were washed with 5 mL of PBS and centrifuged at a maximum of 1500 g for 5 min at RT. A portion of the cell pellet was fixed overnight in 10% formalin in PBS in 1.5 mL microcentrifuge tube. After fixation, samples were briefly centrifuged in a tabletop centrifuge to pellet the cells, the supernatant was removed, and the pellet was resuspended in warm 2% agar at approximately a 1:1 volume ratio. The agar-cell suspension was rapidly mixed and briefly centrifuged to concentrate cells at the base of the agar matrix and then allowed to solidify at 4 °C for at least 5 min. Solidified agar blocks were removed from the tubes, longitudinally bisected, and both halves placed cut-surface down into tissue cassettes containing filter paper. The cassette was then placed in a magnus tissue processor and processed to paraffin according to standard procedures. After processing the halves are placed into a mold, cut surface facing downward. Hot paraffin is added to the mold with the cassette. After cooling, the paraffin block is removed from the mold and then paraffin excess is removed.

This study has been approved by the institutional review boards (study number CFMPB411) of the Netherlands Cancer Institute and of the Portuguese Oncology Institute of Porto, Portugal. Informed written consent was provided by all participants in the study. The research conducted adhered to all relevant guidelines and regulations. The research methods adhered to the principles outlined by the Declaration of Helsinki.

### TMA generation

Tissue Micro Arrays (TMA’s) are made automated using the TMA GrandMaster (3D histech, Sysmex). HE’s were annotated digitally using SlideScore. The annotated images are imported in the GrandMaster Software and overlaid over the picture of FFPE block, so the region of interest for TMA can be selected. For Xenium series of TMAs, cores of 1mm diameter are used, the total array size is 7×13 cores. For orientation purposes 1 core containing healthy spleen was added.

### Xenium Slides Preparation and Run

Xenium tissue slides were prepared by FFPE tissue sectioning at 5 µm thickness and section placement on the Xenium slides according to manufacturer’s protocol (CG000578, 10x Genomics), followed by deparaffinization & decrosslinking according to manufacturer’s protocol (CG000580, 10x Genomics).

The predesigned Xenium Prime 5K Human Pan Tissue & Pathways Panel was used to prepare the tissue samples for Xenium Prime In Situ Gene Expression analysis. Priming probe hybridization, RNase treatment & polishing, 5K Human Pan Tissue & Pathways probe hybridization, ligation and rolling circle amplification were performed. Next, cell segmentation staining reagents were used for labeling of cell nuclei, membranes and interiors to enable automated morphology-based cell segmentation analysis. As the final step in preparing these tissues, the autofluorescence was quenched. All above steps are according to the manufacturer’s protocol (CG000760, 10x Genomics).

Xenium tissue slides were loaded into the Xenium Analyzer Instrument (10x Genomics) for imaging and signal decoding, according to the manufacturer’s protocol (CG000584, 10x Genomics).

Finally Post-Xenium Analyzer H&E Staining was performed according to the manufacturer’s protocol (CG000613). The image acquisition was performed using a Plan-Apochromat 20x/0.8 M27 objective (Zeiss AxioScan Z1) with 0.22 µm/pixel resolution.

### Spatial transcriptomics data analyses

#### Spatial RNA profiling

TMA slides (TMA13 together with TMA15, and TMA17 in a separate run) were analyzed in a 10x Genomics Xenium Analyzer software (v3.1.1.2), using the 5001 gene Xenium Human 5K Pan Tissue & Pathways Panel (v1.0.0). Cell segmentation was performed on the device using the 4-channel multi-modal cell staining. *QuPath* (84) (v0.5.1) (RRID:SCR_018257) was used to create the TMA grid on the multi-color cell morphology images. Cell locations were imported into QuPath and assigned to TMA cores using a custom script.

#### Spatial Xenium data processing

All data analysis was performed in R version 4.4.2 (85) (RRID:SCR_001905) using *tidyverse* (RRID:SCR_019186), *ggplot2* (RRID:SCR_014601), *ComplexHeatmap* (86) (RRID:SCR_017270), *ggalluvial* (87) (RRID:SCR_021253). Spatial cell locations and gene expression data were imported in R using *Seurat* (88) (RRID:SCR_016341) (v5.2.1). In addition to the full slide data the TMA was split into separate objects for each donor block represented by two cores and serialized to disk. Prior to any analysis step the required data is read and combined, filtered using a minimum of 40 transcripts from at least 15 unique genes per cell, normalized to a total of 100 transcript counts per cell and log1p-transformed. UMAP projection and cell clustering is performed using *Seurat* (88).

#### TMA core heterogeneity

Replicate core heterogeneity was estimated for each replicated core by processing the cells as described above. Heterogeneity score is calculated from the normalized minimum contribution by a core for each detected cluster. The final score is the weighted average of the minimum fractions, where the weight is the number of cells in each cluster. For further analyses, we considered the two cores as an unique pooled entity.

#### Cell type labeling

Cell identities were transferred from a single cell experiment performed on a comparable cell type (21) using the RCTD function in the *spacexr* (33) package (v2.2.1). The reference set has labels for a major and a minor cell type that were both transferred in separate runs. The first reported cell label was used as the cell identity.

#### Copy number inference

Data was grouped per patient and processed using *InferCNV* (57) (v1.22) (RRID:SCR_021140) using an adjusted gene filter cutoff of 0.02 to compensate for the lower number of transcripts in Xenium-generated spatial data. The copy number for the cells with either a normal or cancer epithelial cell-type was inferred using all other cell types as the reference set. From the resulting de-noised gene to cell copy number matrix an additional Seurat Assay was created and clustered using the Louvain algorithm (resolution: 0.5). The resulting CNV clusters are used as metadata for subsequent visualization and CNV cluster differential gene-expression analysis.

#### Differential gene expression

All differential gene expression were performed using *Seurat*’s FindMarkers using the Mann-Whitney U-test for the samples with a minimum of 50 tumor cells per sample. Genes were considered differentially expressed when Bonferroni adjusted p-values < 0.01. For inter-patient comparisons, the differentially expressed genes were filtered using a minimum of 30% of the cells (min.pct > 0.3) expressing the gene and a minimum absolute fold-change of 2 (|log₂(Fold Change)| > 1). Whereas, for intra-patient comparisons the resulting gene expression tables were filtered for genes with a |log₂(Fold Change)| > 0.5, a minimum absolute difference in the fraction of cells expressing the gene (|pct.2 − pct.1| > 0.1) or both groups expressing the cells in at least 20% of the cells (pct.1 > 0.2 & pct.2 > 0.2).

#### Pseudotime trajectories definition

Cancer cells derived from individual patients were imported in *Seurat* (88) (v5.3.0) and analyzed independently. *Seurat* FindNeighbors (PCA dims: 1-to-50) followed by FindClusters (resolution: 1.2; random.seed: 42; algorithm: 2) was used to define cell clusters by gene expression. Resulting Seurat object was converted in a singleCellExperiment object and used to infer pseudotime using *slingshot*(61) (v2.12.0) (reducedDim: UMAP, maxit: 50; dist.method: mnn).

To define the genes that most contributed to each trajectory, slingPseudotime and slingCurveWeights, from *slingshot*, were used to extract pseudotime and membership scores to a given trajectory per each cell, respectively. Only cells with at least one assigned trajectory were retained. Then, smoothers along pseudotime were fitted gene-wise for a given trajectory to Generalized Additive Model (GAM) using fitGAM (counts: log-normalized counts, nknots: 6, family: gaussian – to take into account the distribution of log_2_-normalized counts –) from *tradeSeq* (62) (v1.18.0). Genes have been then tested for their association to pseudotime using associationTest from *tradeSeq* and adjusting the p-value using the Benjamini-Hochberg (BH) method. The predicted smooth for each gene is then extracted using predictSmooth from *tradeSeq* in order to compute a trajectory-specific effect size from the fitted curves. Then, we computed the effect relative to the maximum effect identified (effect_range: max(yhat) - min(yhat); where the ‘yhat’ is the fitted expression). Per each trajectory, each gene was then ranked by BH-adjusted p-value and effect_range; top 50 genes for each trajectory were then selected for further analyses.

### Over-representation analyses (ORA)

ORA analyses were performed using *clusterProfiler* (89) (v4.12.6) on query sets of genes. Enrichment was computed over the signatures of Hallmarks (59) and estrogen response curated by Li *et al.*(60). The original signatures have been filtered to contain only genes available in the 5000 genes quantifiable by the Xenium spatial transcriptomics platform (10x Genomics). Only signatures for which the enrichment associated q-value was lower than 0.05 were retained.

### Downstream statistical analysis and data visualization

Statistical analyses and data visualizations of processed data were performed in R (85) (v4.4.2), with figures generated using *ggplot2* (v4.0.1; RRID:SCR_014601) and *pheatmap* (v1.0.13, RRID:SCR_016418).

### Immunohistochemistry

Immunohistochemistry of the FFPE tumor samples was performed on a BenchMark Ultra autostainer (Ventana Medical Systems). Briefly, paraffin sections were cut at 3 μm, heated at 75°C for 28 min and deparaffinized in the instrument with EZ prep solution (Ventana Medical Systems). Heat-induced antigen retrieval was carried out using Cell Conditioning 1 (CC1, Ventana Medical Systems) for 36 min at 95°C (ERα, PR, HER2, p-S6 Ribosomal Protein (Ser235/236), E-Cadherin).

ERα was detected using clone SP1 (Ready-to-Use, 32 min at RT, #5278406001, Roche Diagnostics/Ventana), PR using clone 1E2 (Ready-to-Use, 32 min at RT, #5277990001, Roche Diagnostics/Ventana), HER2 clone 4B5 (Ready-to-Use, 12 min at RT, #5278368001, Roche Diagnostics/Ventana), Ki-67 clone 30-9 (Ready-to-Use, 32 min at 37°C, #5278384001, Roche Diagnostics),Phospho-S6 Ribosomal Protein (Ser235/236) Polyclonal (1:50 dilution, 1 hour at RT, #2211S, Cell Signaling). To reduce background signal for the PR staining, after primary antibody incubation slides were incubated with normal antibody diluent (Roche Diagnostics) for 24 min.

Bound antibody was detected using the UltraView Universal DAB Detection Kit (Ventana Medical Systems) for ERα, PR, HER2, p-S6 Ribosomal Protein (Ser235/236). Slides were counterstained with Hematoxylin and Bluing Reagent (Ventana Medical Systems).

A PANNORAMIC^®^ 1000 scanner from 3DHISTECH was used to scan the slides at a 40x magnification. The scoring of the samples was done using the online platform Slidescore 500 (www.slidescore.com).

### Automated multiplex staining on Discovery Ultra Stainer and HALO imaging

Prior to multiplex staining 3µm slides were cut on TOMO slides. Slides were then dried overnight and stored in +4°C. Before a run was started slides were baked for 30 min at 70°C in an oven.

Staining was performed on a Ventana Discovery Ultra automated stainer, using the Opal 6-Plex Detection Kit (50 slides kit, Akoya Biosciences, NEL871001KT). Protocol starts with baking for 28 min at 75°C, followed by dewaxing with Discovery Wash using the standard setting of 3 cycles of 8 min at 69°C. Pretreatment was performed with Discovery CC1 buffer for 64 min at 95°C, after which Discovery Inhibitor was applied for 8 min to block endogenous peroxidase activity. Specific markers were detected consecutively on the same slide with the following antibodies, Anti-CD68 (Clone KP1, M0814, DAKO, 1/200 dilution 1h at RT), anti-CD8 (Clone C8/144B, M7103, DAKO, 1/50 dilution, 1h at RT), anti-FoxP3 (clone 236A/E7, ab20034, Abcam, 1/100 dilution, 2h at RT), anti-CD3 (Clone SP7, RM-9107-S,Thermo Scientific, 1/200 dilution 1h at RT), Anti-PanCK (Clone AE1/AE3, MS343P, Thermo Scientific, 1/100 dilution, 2h at RT) and Anti-CD20 (Clone L26, M0755, DAKO, 1/500 dilution, 1h at RT). Each staining cycle was composed of four steps: Primary Antibody incubation, Opal polymer HRP Ms+Rb secondary antibody incubated for 1h at RT, OPAL dye incubation (OPAL480, OPAL520, OPAL570, OPAL620, OPAL690, OPAL780, 1/40 or 1/50 dilution as appropriate for 1 hour at RT) and an antibody denaturation step using CC2 buffer for 20 min at 95°C. Cycles were repeated for each new antibody to be stained. At the end of the protocol slides were incubated with DAPI (1/25 dilution in Reaction Buffer) for 12 min.

After the run was finished slides were washed with demi water and mounted with Fluoromount-G (Southern Biotech, 0100-01) mounting medium.

After staining slides were imaged using the PhenoImager HT automated imaging system (Akoya). Scans were made with the MOTiF protocol. Using the InForm software version 2.5.0 the MOTIF images were unmixed into 8 channels: DAPI, OPAL480, OPAL520, OPAL570, OPAL620, OPAL690, OPAL780 and Auto Fluorescence and exported to a multilayered TIFF file.

The HALO software (V4.0.5107.357, Indica Labs) was used for image-analysis. Tissue Microarray cores were selected using the annotation tool. The Indica Labs HighPlex FL v4.2.14 analysis algorithm was used for analysis using AI nuclei segmentation (Nuclei Seg V2; HALO AI). ROI’s from three technical replicates (cores) were pulled together and analyzed. Both the summary data and cell object data were exported in comma separated value files using the export manager in HALO.

### Spatial Flow Cytometry (SFC)

Cells were counted and seeded in triplicate at 30,000 live cells per well in a 96-well U-bottom plate 3799, Corning). Fluorescent counting beads (100 µL per well; 1:25; ACFP-50-5, Spherotech) were added prior to centrifugation. Cells and beads were pelleted at 300 x g for 5 min at 4 °C, after which the supernatant was removed. Pellets were washed twice with PBS, followed by centrifugation at 300 x g for 5 min at RT. Viability staining was performed by incubating cells with LIVE/DEAD™ Fixable Near-IR (780) viability dye (200 µL per well; 1:1000; L34993, Invitrogen) for 15 min at RT, protected from light. Samples were washed with cell staining buffer (420201, BioLegend) and incubated with Human Fc Block reagent (50 µL per well; 1:1000; 564219, BD Biosciences) for 10 min at RT. Subsequently, cells were stained with a phenotyping antibody cocktail in cell staining buffer (50 µL per well; CD45-BUV395, 563792, BD Biosciences; CD8-BUV496, 612943, BD Biosciences; CD56-BV421, 362552, BioLegend; CD15-BV510, 323028, BioLegend; EpCAM-BV605, 324224, BioLegend; CD4-BV785, 317441,

BioLegend; CD11b-PE, 301306, BioLegend; CD3-PE-Dazzle594, 317346, BioLegend; CD71-PE-Cy7, 334111, BioLegend; FAPa-AF647, FAB3715R, R&D Systems; CD19-APC-FIRE750, 302257, BioLegend) for 30 min at 4°C in the dark; unstained control wells received cell-staining buffer only. Following staining, samples were washed, resuspended in 50 µL of cell staining buffer, and acquired on a 5-laser spectral flow cytometer (ID7000, Sony Biotechnology).

Spectral unmixing was performed using instrument-specific software (Spectral Analyzer ID7000, Sony Biotechnology), using reference spectra generated for each fluorochrome included in the panel. Autofluorescence was identified as an independent spectral component and subtracted during unmixing. Unmixed flow cytometry data were analyzed using FlowJo (version 10.10, BD Biosciences) and Cytolution (Cytolytics). Doublets, debris, and beads were excluded prior to conventional phenotypic analysis. Finally, to enable quantitative comparison between samples, cell counts were normalized to fluorescent counting beads included in each well at a fixed concentration prior to acquisition.

### MicroOganoSpheres (MOS) preparation and treatment

Fresh-frozen MPE samples were thawed and incubated with DNAse-I (0.001 U/µL; EN0521, Thermo Scientific) for 10 mins, followed by Accutase (2868335, Gibco) for 5 min. Subsequently, MicroOrganoSpheres (MOS) were generated as previously described (54,55). MOS were resuspended in culture medium (adapted from Sachs *et al.* (90), plated in 96-well PhenoPlate (6055302, Revvity) and cultured for 4 days in dimethyl-sulfoxide (DMSO; D8418-50ML, Sigma-Aldrich), 50µM Carboplatin (HY-17393, MedChemExpress), or 1µM or 10µM Tamoxifen (HY-16950, MedChemExpress) conditions. At the start and end-point of the assay, prior to staining for SFC (see “*Spatial Flow Cytometry*” section in the *Material and Methods*), MOS were dissociated through incubation with Dispase II (17105041, Gibco) for 30 min at 37°C, followed by 5 min incubation with Accutase.

