## Supplementary Figure for "A spatial single-cell transcriptomic atlas of metastatic breast cancer progression"

### Supplementary Figure 1

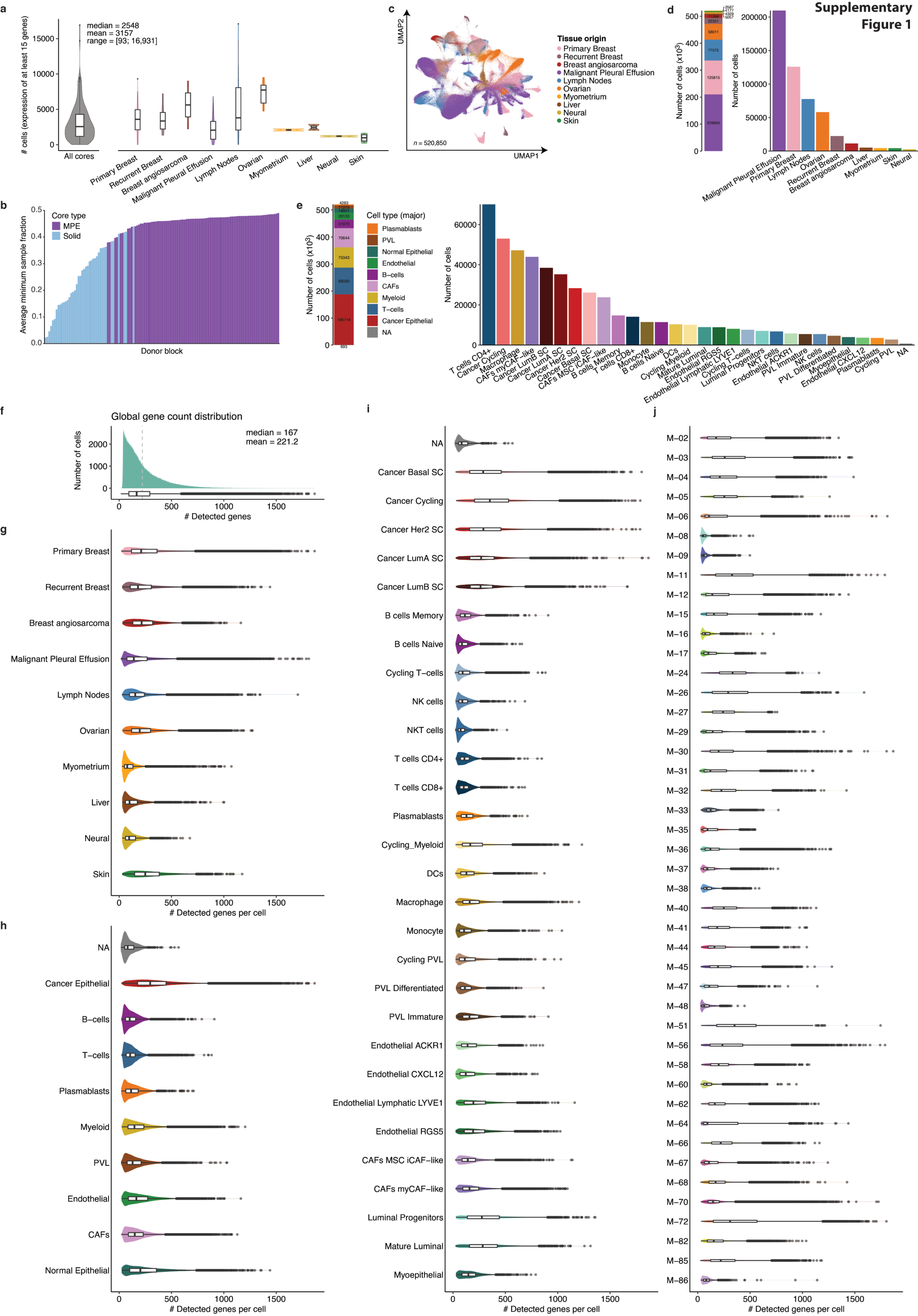

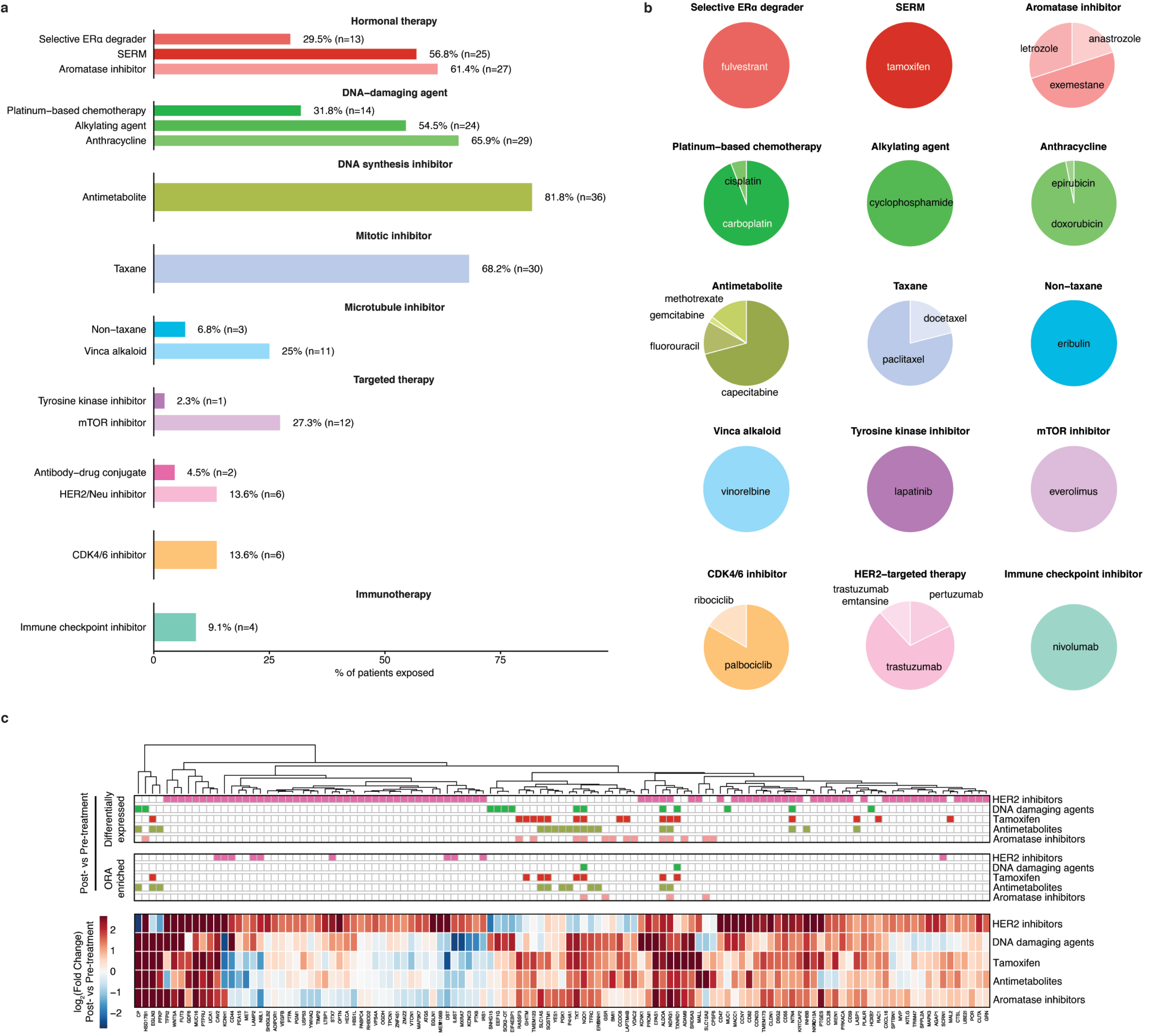


Supplementary Figure 2

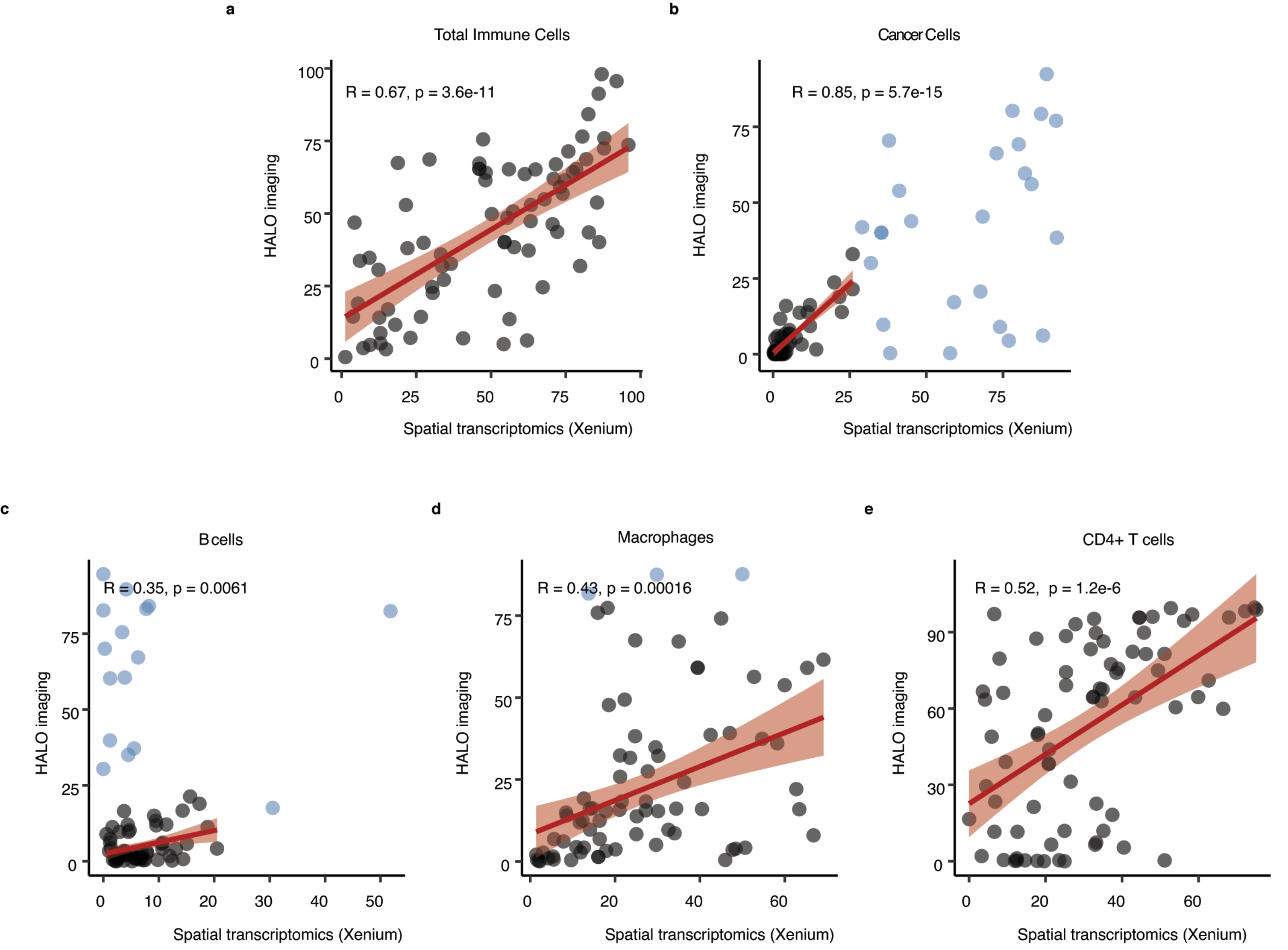


Supplementary Figure 3

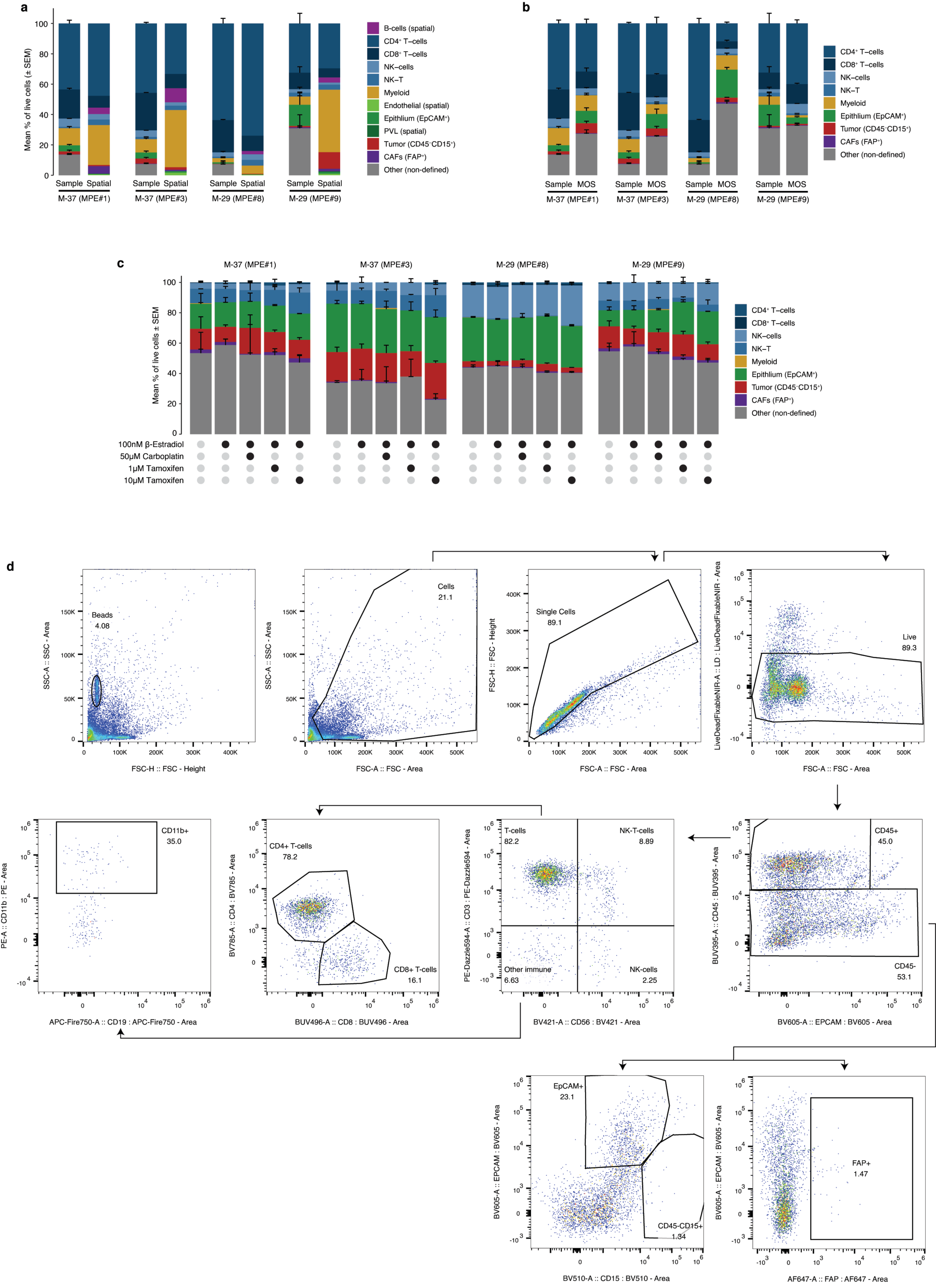


Supplementary Figure 4

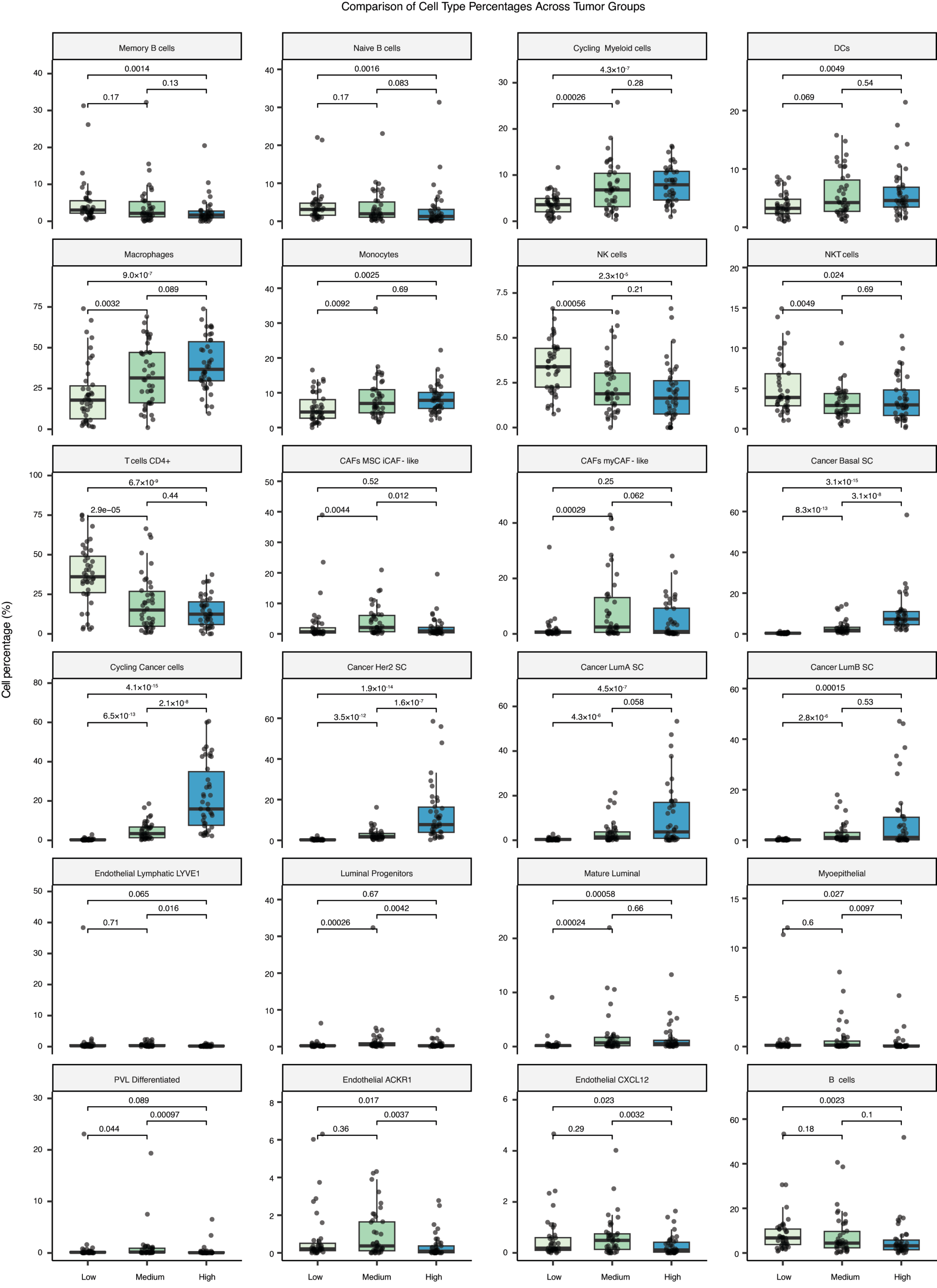


Supplementary Figure 5

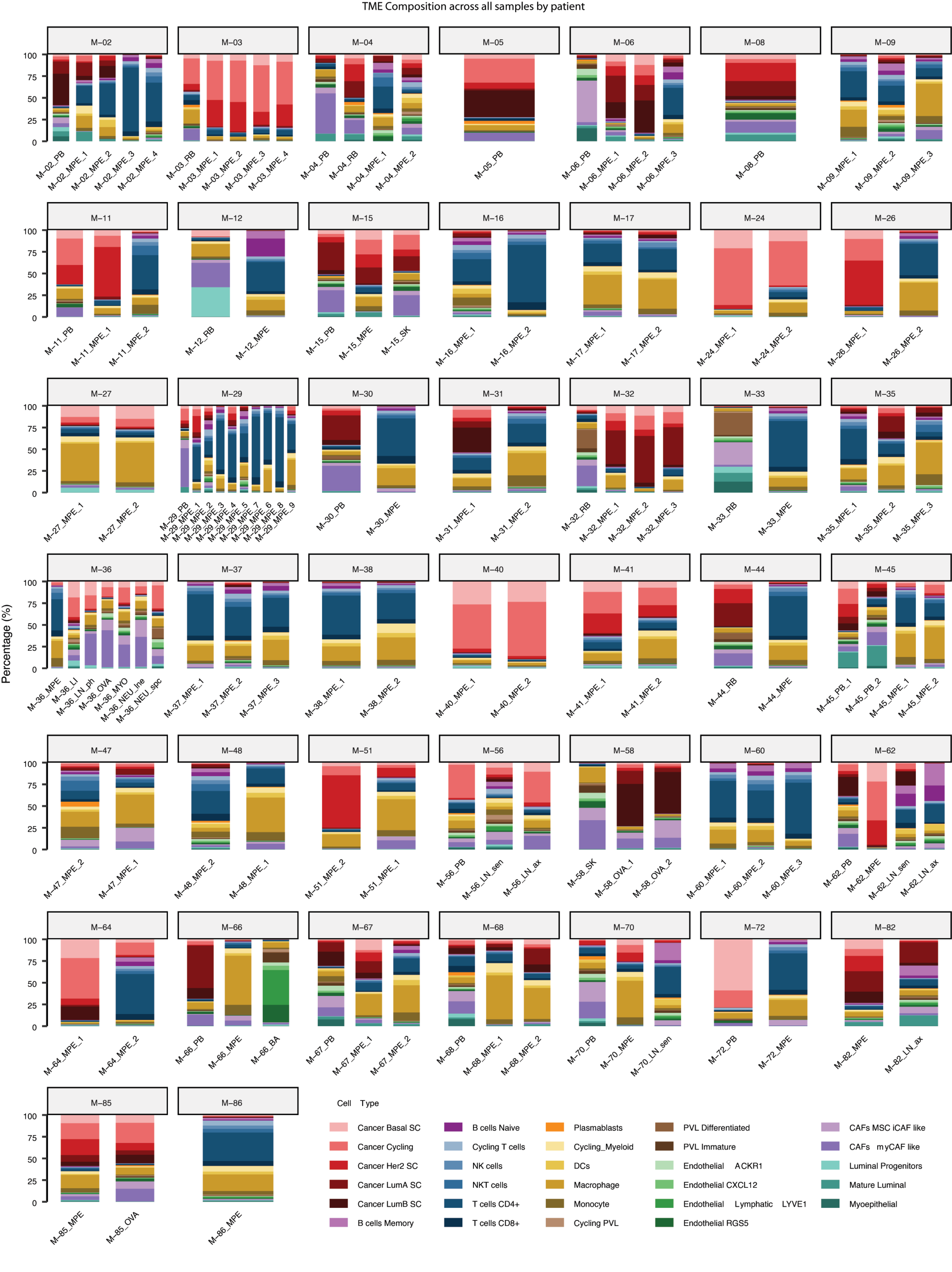


Supplementary Figure 6

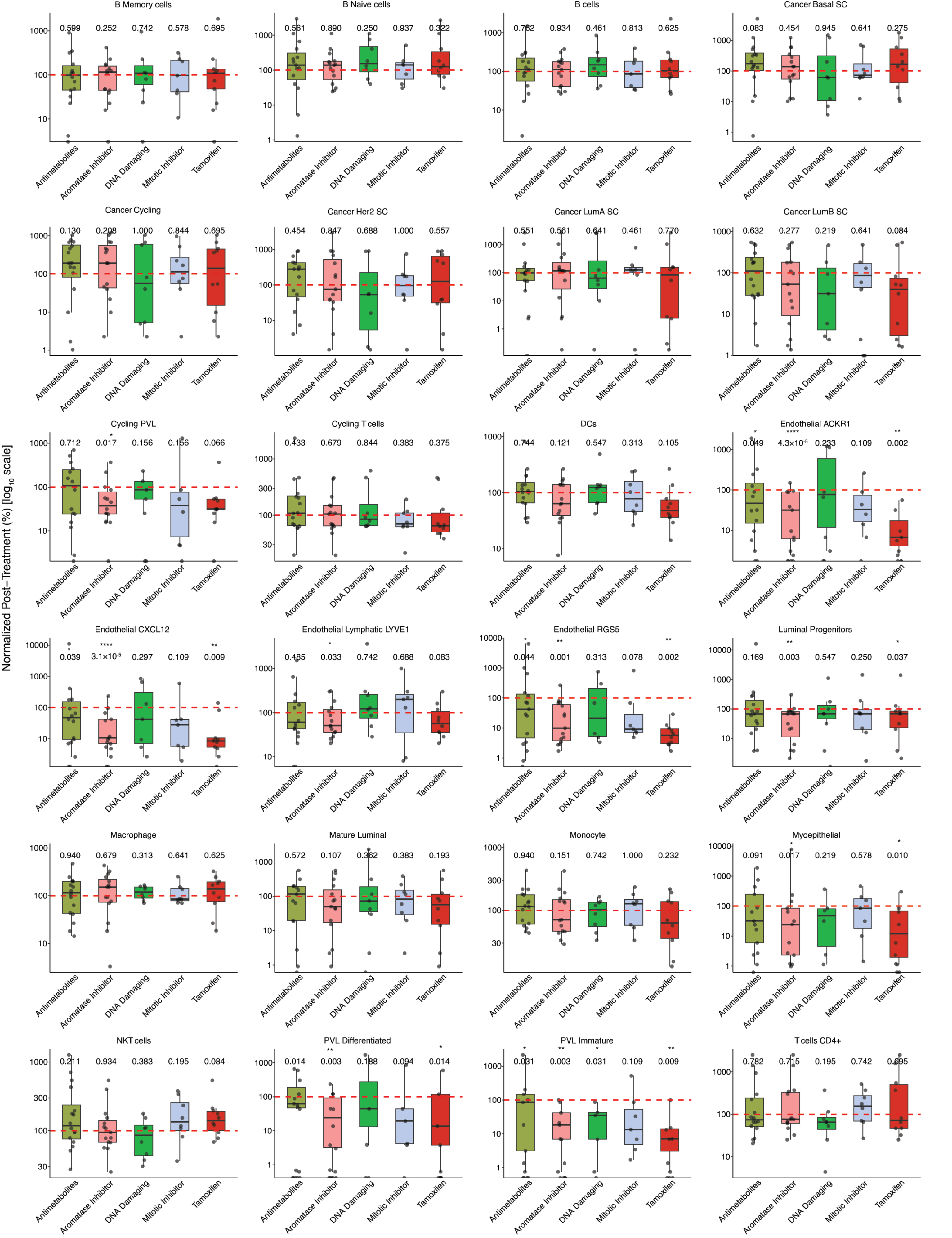


Supplementary Figure 7

Supplementary Figure 8

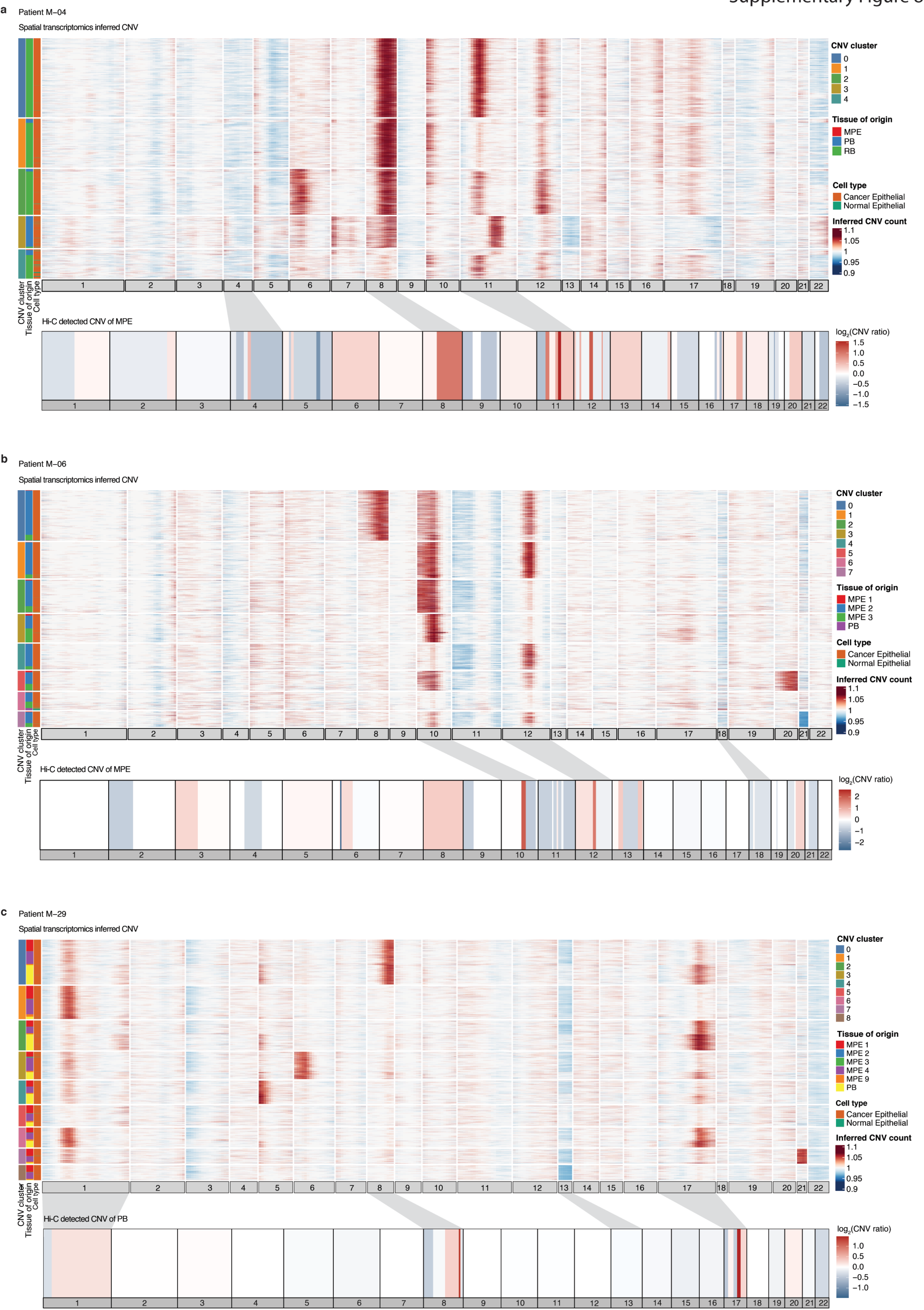

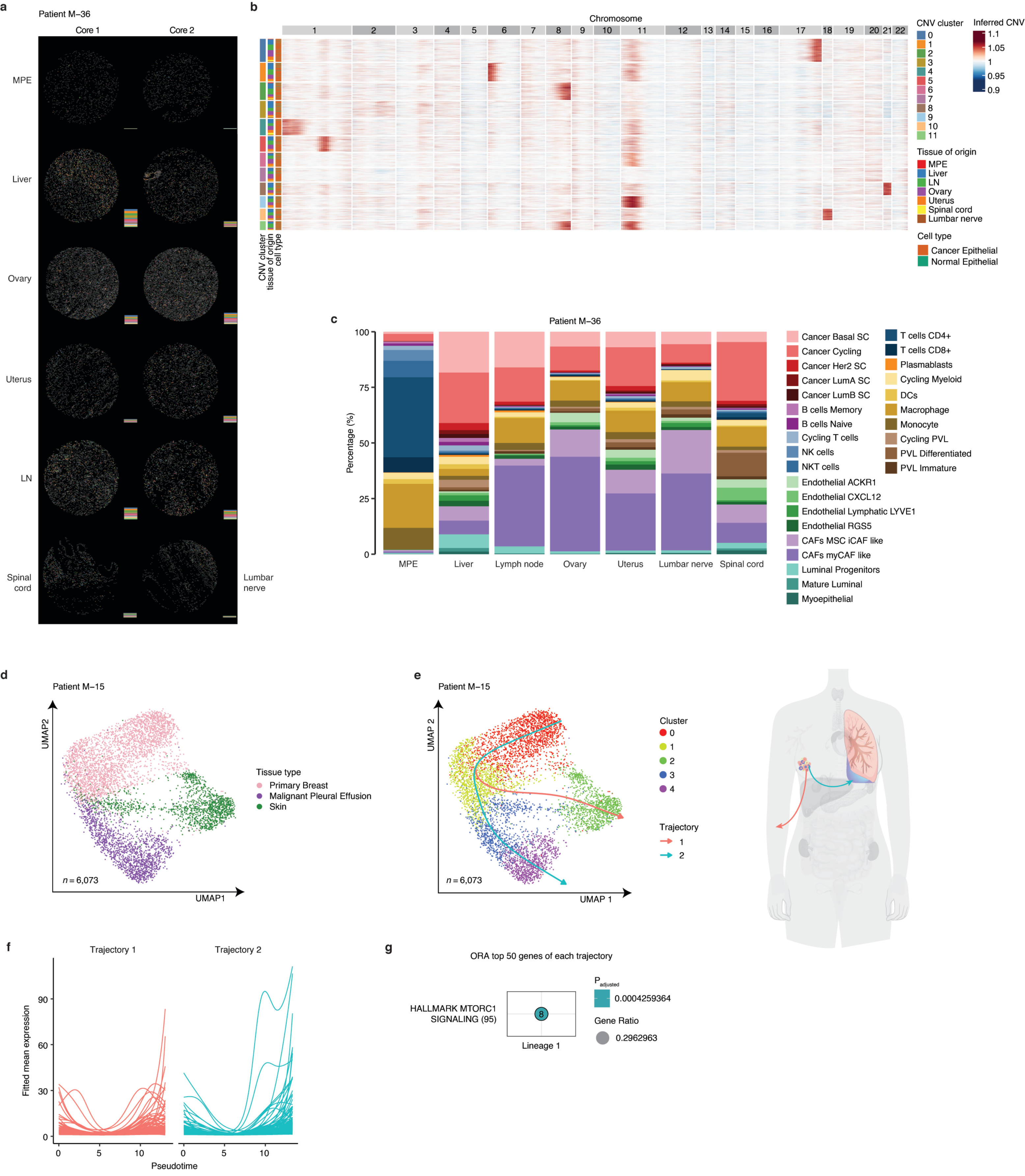


Supplementary Figure 9
